# Integrating sequence-to-omics predictions and personal multi-omics for rare variant prioritization

**DOI:** 10.64898/2026.10.24.26366361

**Authors:** Julie Lake, Esther Robb, Victoria Rosa, Taylor M. Arriaga, Rebekah Barrick, Seth I. Berger, Benjamin D. Heavner, Charles Hadley King, Arthur Ko, Jonathan LoTempio, Daniel Nachun, Genomics Research to Elucidate the Genetics of Rare Diseases consortium, Anshul Kundaje, Devon E. Bonner, Emmanuèle C. Délot, Eric Vilain, Matthew T. Wheeler, Stephen B. Montgomery

## Abstract

Sequence-to-omics (S2O) predictions and molecular outliers provide complementary evidence to prioritize functional rare variants and facilitate rare disease diagnoses. However, current approaches do not integrate both lines of evidence. We present Watershed-DL, a framework that integrates S2O predictions of variant effects from DNA and protein sequence with multi-omic outliers to identify large-effect rare variants. We applied Watershed-DL to 1000 Genomes samples with RNA-seq (n=1,030) to prioritize rare variants underlying expression, splicing, and allele-specific expression outliers, and, using a separate peak-level model, to samples with ATAC-seq (n=174) for chromatin accessibility outliers. Compared to a model without S2O predictions, Watershed-DL increased the number of prioritized variants by 61.4%, and S2O features were weighted highly in the model across all molecular signals. Prioritized variants were over 60-fold enriched for ClinVar pathogenic classifications. Matching prioritized variants to known pathogenic variants in the same gene with a concordant molecular effect identified 331 additional candidate recessive disease alleles. Applying this framework to the GREGoR rare disease cohort with RNA-seq (n=677) prioritized previously reported causal variants across 10 solved cases and identified 451 candidate variants in unsolved cases, including a diagnostic variant in *PPP1R12A* and a novel candidate disease gene, *ATP5PF*.

---

Rare genetic variants are abundant in human populations, yet their functional effects remain difficult to characterize ^1^. While large-scale biobank studies have shown that rare variants contribute to heritability across numerous traits ^2,3^, their individual effect sizes are challenging to estimate due to limited statistical power. As population-scale whole genome sequencing efforts continue to identify hundreds of millions of rare variants, scalable approaches for interpreting their functional effects are needed to improve their identification and use in rare disease diagnosis and precision health.

Recent advances in deep learning have enabled genome-wide prediction of variant functional effects directly from DNA and protein sequence. Sequence-to-omics (S2O) models can now predict the effects of genetic variation across the allele frequency spectrum for diverse molecular phenotypes, such as gene expression ^4–6^, chromatin accessibility ^7^, splicing ^8,9^, polyadenylation ^10^, and protein function ^11–13^. While these approaches facilitate functional scoring of variants, they have shown limitations in capturing individual variability of molecular signals ^14^ and provide only indirect evidence of variant function. At the same time, the growing availability of transcriptomic and other functional genomic data from large population and disease cohorts provides new opportunities to characterize the functional effects of rare variants by integrating S2O predictive models with individual multi-omics.

Complementing S2O predictions, extreme or outlier deviations in molecular phenotypes can provide evidence of functional effects of nearby rare variants ^15–17^. These molecular outliers have proven valuable for rare disease diagnosis and the interpretation of candidate genetic variants ^18–20^. However, systematically linking outlier signals to their underlying genetic variants remains challenging at scale. Molecular outlier calling methods typically detect aberrant phenotypes first and subsequently use variant annotation or manual evaluation to identify candidate variants, rather than jointly modeling both signals ^21,22^. We previously designed RIVER ^16^ to address this challenge by integrating molecular outliers with variant-level annotations, and Watershed ^15,23–25^ to extend this approach by combining evidence across multiple molecular phenotypes. Despite these advances, neither approach has incorporated information about specific regulatory or protein pathogenicity effects of variants prioritized by S2O predictive models.

To address these limitations, we developed Watershed-DL, a framework that integrates S2O predictions with multi-omic outliers for rare variant prioritization. By leveraging complementary signals, Watershed-DL jointly models predicted and observed molecular effects to improve the functional interpretation of rare variants. We evaluated the performance of Watershed-DL in the 1000 Genomes Project (1KG), and demonstrated that integrating multi-omics with S2O predictions improved functional variant discovery compared to using either data source independently. We applied this framework alongside molecular effect matching to prioritize candidate variants in recessive disease genes carried by individuals in the 1KG, including variants in non-coding regions and in ancestry groups underrepresented in ClinVar. We then applied this framework to solved and unsolved rare disease patients from the Genomics Research to Elucidate the Genetics of Rare Diseases (GREGoR) Consortium ^26^. Watershed-DL prioritized a homozygous splice variant in a candidate novel disease gene *ATP5PF* in one unsolved proband and, in another, independently identified a previously reviewed *PPP1R12A* missense variant, providing the mechanistic evidence needed for a diagnosis.

## Results

### Multi-omic outliers and S2O predictions provide complementary evidence of rare variant function

Watershed-DL integrates S2O predictions and other variant-level annotations alongside personal multi-omic outlier signals in a probabilistic graphical model to predict the functional effects of rare variants (Figure 1A). The design of Watershed-DL allows for information sharing across different multi-omics data types and variant-level annotations. We integrated S2O predictions as variant annotations in Watershed-DL, including models of gene expression (Flashzoi-RNA ^27^, PromoterAI ^6^), alternative splicing (SpliceAI ^8^, Pangolin ^9^), alternative polyadenylation (APARENT2 ^10^), chromatin accessibility (ChromBPNet ^7^), and protein structure and function (ESM-v1 ^11^, PrimateAI ^12^, AlphaMissense ^13^). We also develop a novel extension of the Watershed-DL framework for chromatin accessibility outliers, included as a separate model, to prioritize rare variants within ATAC-seq peaks.

**Figure 1:**
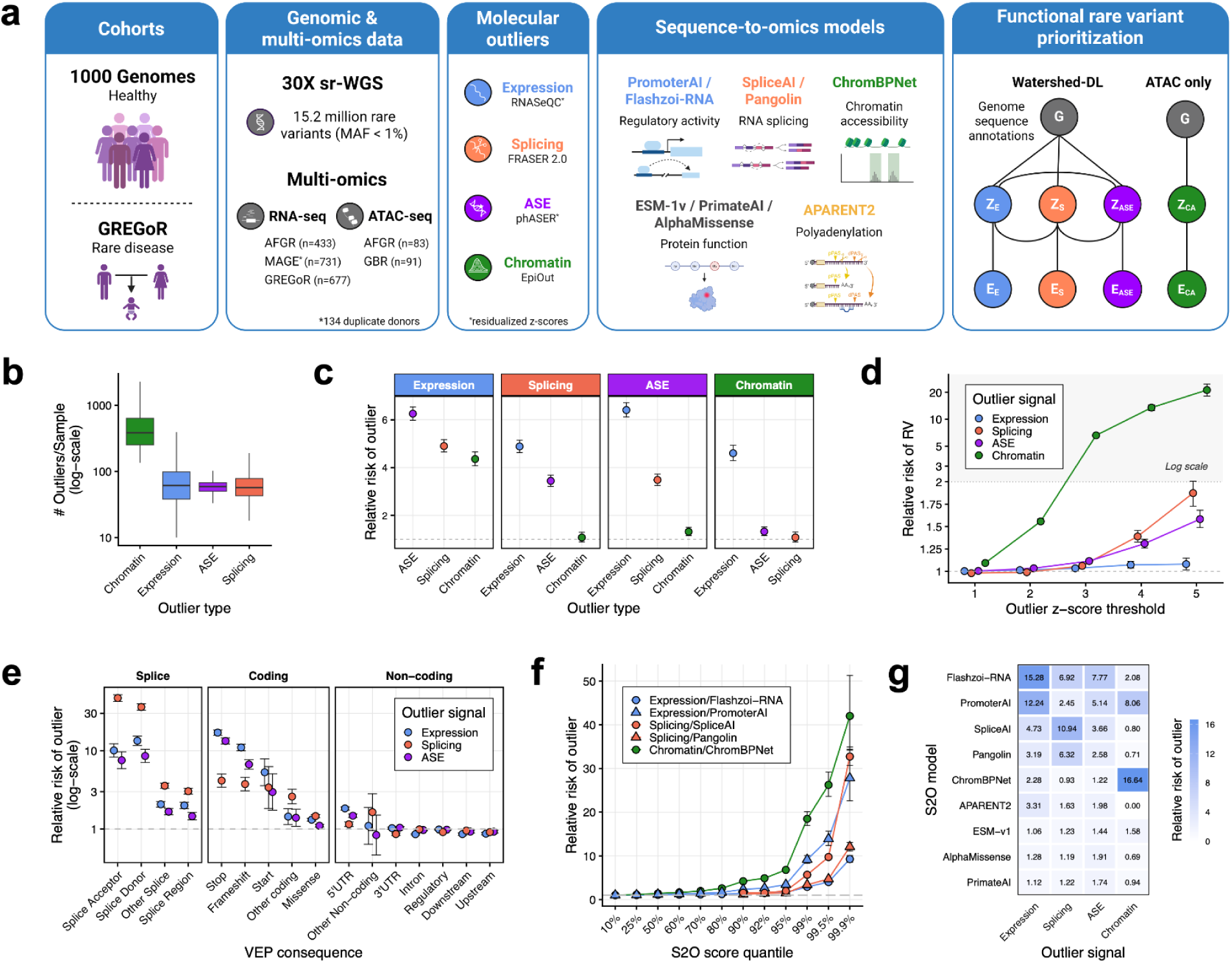
Multi-omics outlier discovery and rare variant functional stratification. a) Study design and Watershed-DL framework. Molecular outliers were identified in 1KG and GREGoR samples with 30X sr-WGS and matched RNA-seq and/or ATAC-seq, and rare variants (RVs) were annotated with nine S2O models. b) Molecular outliers per individual in the 1KG, by outlier signal. c) Pairwise enrichment of outlier signals in the 1KG, shown as the relative risk (RR) of an outlier in one molecular phenotype given an outlier in another. Transcriptomic outliers were identified in 1,030 donors with sr-WGS and RNA-seq, and caOutliers in 111 donors with sr-WGS, RNA-seq, and ATAC-seq. d) RR of RVs within 10 kb of genes with eOutliers, aseOutliers, or sOutliers, or within caOutlier peaks, across |z| thresholds. e) RR of RVs by Ensembl Variant Effect Predictor (VEP) consequence across outlier signals. Outliers were assigned to the most consequential nearby rare variant. caOutliers were excluded as variants are not annotated with respect to a specific gene. f) RR of an outlier among RVs with S2O predictions above the indicated quantile, for each molecular phenotype and its matched model. g) Enrichment of S2O-prioritized RVs across outlier signals. Thresholds use absolute scores unless noted: APARENT2 ≥ 1.45, ChromBPNet ≥ 0.5, ESM-1v ≤ −7.5 (signed), Flashzoi (RNA) ≥ 0.2, Pangolin ≥ 0.2, PrimateAI ≥ 0.8, PromoterAI ≥ 0.5, SpliceAI ≥ 0.2, and AlphaMissense ≥ 0.564 (likely pathogenic).

We applied Watershed-DL to lymphoblastoid cell line (LCL) RNA-seq data from 1,030 donors from the 1KG generated as part of AFGR ^28^, representing 5 African populations, and MAGE ^29^, representing 26 global populations. From these samples, we identified gene-level outliers for RNA expression (eOutliers), allele-specific expression (aseOutliers), and alternative splicing (sOutliers) (see Methods). For the peak-level adaptation of Watershed-DL, we identified chromatin accessibility outliers (caOutliers) from LCL-derived ATAC-seq data from 174 samples, generated by AFGR ^28^ and by Kumasaka *et al.* (British (GBR) individuals) ^30^. For each outlier signal, z-scores were corrected for batch-driven sample covariation (Figure S1; see Methods). After quality filtering, 14,312 genes were quantified for eOutlier identification, 12,740 for sOutliers, and 10,294 for aseOutliers, while 194,385 peaks were jointly called for caOutlier identification. We then applied Watershed-DL to 15.2 million rare variants (global maximum allele frequency < 1% in gnomAD v4.1) identified from 30X short-read whole genome sequencing (sr-WGS) of 1KG donors with matched RNA-seq or ATAC-seq (Figure S2).

We characterized the distribution and overlap of molecular outliers across individuals. Chromatin accessibility outliers were more abundant per individual than other molecular phenotypes measured at the gene level (Figure 1B), consistent with the greater number of accessibility peaks compared to genes. We observed a median of 383 caOutliers (interquartile range (IQR), 248-624), compared with 61 eOutliers (IQR, 38-98), 59 aseOutliers (IQR, 50-67), and 57 sOutliers (IQR, 43-78). Outliers were frequently shared across molecular phenotypes, with the strongest co-occurrence between eOutliers and aseOutliers (relative risk (RR) = 6.41), followed by eOutliers and sOutliers (RR = 4.90), eOutliers and caOutliers within 10 kilobases (RR = 4.60), and sOutliers and aseOutliers (RR = 3.48) (Figure 1C).

We first tested whether molecular outliers were associated with an increased burden of nearby rare variants. Without incorporating functional annotations, rare variants were enriched within 10 kb of genes with eOutliers, aseOutliers, and sOutliers, as well as within caOutlier peaks, with enrichment increasing with outlier strength (Figure 1D). At an absolute z-score > 3, caOutliers showed the strongest enrichment (RR = 6.61, P < 0.001), followed by aseOutliers (RR = 1.09, P = 2.18 × 10^−26^), sOutliers (RR = 1.06, P = 4.35 × 10^−10^), and eOutliers (RR = 1.02, P = 3.66 × 10^−5^). The enrichment was further differentiated by variant consequence, with splice and coding variants showing stronger enrichment than non-coding variants (Figure 1E).

To assess whether S2O predictions of rare variants in the 1KG were associated with molecular outliers, we evaluated the enrichment of model predictions for corresponding outlier signals. At the 99th percentile of each S2O model, we observed the strongest enrichment of ChromBPNet for caOutliers (RR = 18.48), followed by PromoterAI for eOutliers (RR = 9.14), SpliceAI for sOutliers (RR = 5.68), and Pangolin for sOutliers (RR = 3.40) (Figure 1F). S2O predictions consistently showed the highest enrichment for their matched outlier type at their recommended thresholds (Figure 1G). These results support integrating S2O predictions with multi-omic outliers to prioritize functional rare variants with large molecular effects.

### Watershed-DL improves prioritization of large effect non-coding rare variants

To assess whether integrating S2O predictions with molecular outliers improves rare variant prioritization, we trained Watershed-DL in the 1KG and compared its performance with two baselines: a Watershed model without S2O predictions, and a genome annotation model (GAM) which includes S2O predictions but does not integrate molecular outliers (see Methods). Compared to the Watershed baseline, Watershed-DL demonstrated improved precision-recall area under the curve (PR-AUC) across expression (0.028 vs. 0.023 without S2O), splicing (0.054 vs. 0.041), and chromatin accessibility (0.129 vs. 0.055) (Figure 2A). For ASE, which did not have a specifically matched S2O model, Watershed-DL showed comparable performance to the non-S2O model (PR-AUC = 0.013 vs. 0.014). We additionally compared with an S2O annotation-only baseline and found that the Watershed-DL model improved PR-AUC across all outlier signals (Figure S3). These results demonstrate that integrating S2O predictions with molecular outliers improves performance of functional rare variant discovery.

**Figure 2:**
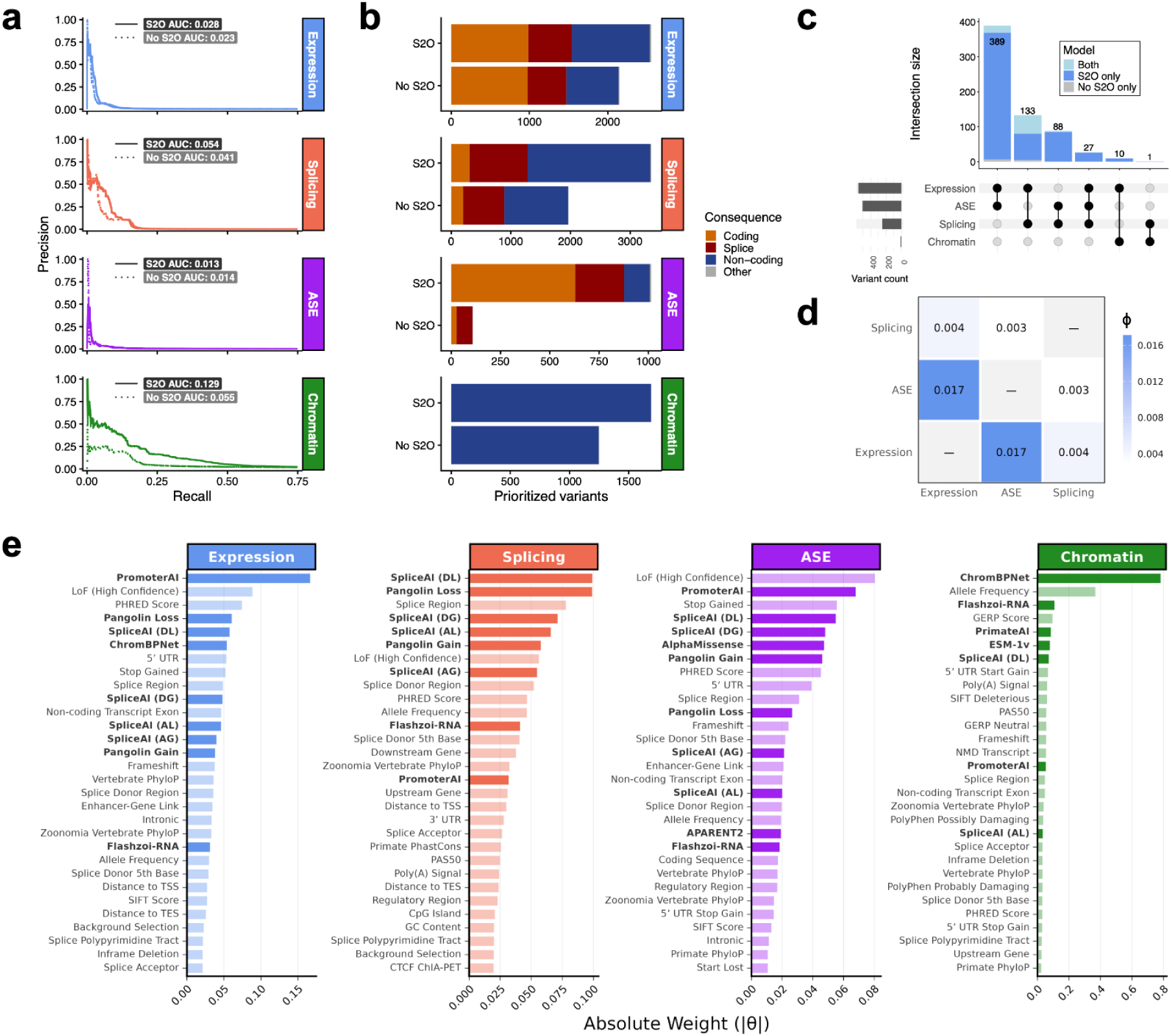
S2O integration and rare variant prioritization performance. a) Precision-recall curves for Watershed-DL (solid lines) compared to Watershed model without S2O predictions (dashed lines). b) Number of prioritized variants for each outlier signal at posterior probability ≥0.6. Prioritized variants were filtered to those with at least one outlier carrier. c) Number of prioritized variants from Watershed-DL that are shared across multiple outlier signals. d) Edge weights (ϕ) connecting latent functional variables (Z) representing shared weights between outlier signals in the Watershed-DL model. e) Edge weights (θ) connecting genomic annotation features (G) to latent regulatory variables (Z) in each outlier signal, ranked by absolute Theta value representing relative informativeness. The top 30 informative annotations are shown for each outlier signal.

In addition, Watershed-DL prioritized more variants across all molecular signals compared to the baseline Watershed model, with the largest increase of prioritized variants in non-coding regions (Figure 2B). At posterior probability ≥0.6, Watershed-DL prioritized 2,555 variants for expression, 3,351 for splicing, 1,014 for ASE, and 1,690 for chromatin accessibility that were associated with at least one outlier carrier. Overall, integrating S2O predictions increased prioritized variants by 61.4% across all outlier signals. Further, 77.9% of newly prioritized variants were neither coding nor canonical splice-site variants, highlighting the ability of S2O predictions to identify functional non-coding variants compared to prior genome annotations based on coding annotations, splice site regions, or conservation metrics. Watershed-DL also prioritized more variants associated with two or more outlier signals compared to the baseline Watershed model. The largest increases were in variants prioritized for both expression and ASE (n = 389), followed by expression and splicing (n = 133), with 86.4% of multi-outlier prioritized variants uniquely identified by Watershed-DL (Figure 2C). Consistent with these overlaps, Watershed-DL learned highest-ranking edge weights connecting the expression and ASE latent variables (θ = 0.017), followed by expression and splicing (θ = 0.004; Figure 2D), indicating that the model leveraged shared functional status across these outlier signals.

Watershed-DL learned consistently high feature weights for S2O predictions across all outlier signals (Figure 2E). Molecular phenotype-matched S2O predictions were the top-weighted features across expression, splicing, and chromatin accessibility, with ChromBPNet as the most informative feature for chromatin accessibility (θ = 0.78), PromoterAI for expression (θ = 0.17), and SpliceAI and Pangolin for splicing (maximum θ = 0.10 for both). For ASE, which did not have a directly matched S2O model, PromoterAI ranked 2nd-most informative (θ = 0.07), and SpliceAI donor gain and loss ranked 4th and 5th respectively (θ = 0.05 for both). In contrast, the Watershed model without S2O predictions weighted allele frequency as the most informative feature for chromatin accessibility (θ = −0.33), loss-of-function classification for expression (θ = 0.10), and splice region annotation for splicing (θ = 0.13) (Figure S4). These findings demonstrate that S2O predictions are highly impactful for prioritizing outlier-associated rare variants across multiple transcriptomic and chromatin accessibility signals, even when compared to high-confidence coding annotations such as loss-of-function.

### Watershed-DL reveals ancestry-specific recessive disease alleles

To evaluate the clinical relevance of prioritized variants in the 1KG, we examined their overlap with established disease gene sets and clinical variant classifications. Watershed-DL identified 1,432 prioritized variants in ClinGen, OMIM, and ACMG recessive disease genes (Figure S5A), providing candidates for further clinical interpretation. ClinVar representation varied across molecular phenotypes (Figure 3A), with ASE variants showing the highest proportion represented in ClinVar (18.41%), followed by expression (13.83%) and splicing (11.12%). Chromatin accessibility variants showed the lowest representation (0.95%), consistent with the under-ascertainment of non-coding regulatory variants in clinical databases ^31^. Among prioritized variants represented in ClinVar (n=801), 166 were previously classified as pathogenic or likely pathogenic (P/LP) and 225 as variants of uncertain significance (VUS). Excluding chromatin accessibility which had very low overlap, pathogenic and likely pathogenic variants were enriched among prioritized variants (RR = 65.20 and RR = 45.41, respectively), whereas benign and likely benign variants were depleted (RR = 0.15 and RR = 0.32, respectively) (Figure 3B). Enrichment for P/LP variants was comparable between the baseline Watershed model and Watershed-DL, consistent with the greater representation of coding and canonical splice-site variants among ClinVar P/LP variants, which are well captured by standard annotations ^32^. S2O integration instead expanded the set of prioritized non-coding and non-canonical splice-site variants, which remain underrepresented in ClinVar but comprised 70.35% of variants prioritized by Watershed-DL.

**Figure 3:**
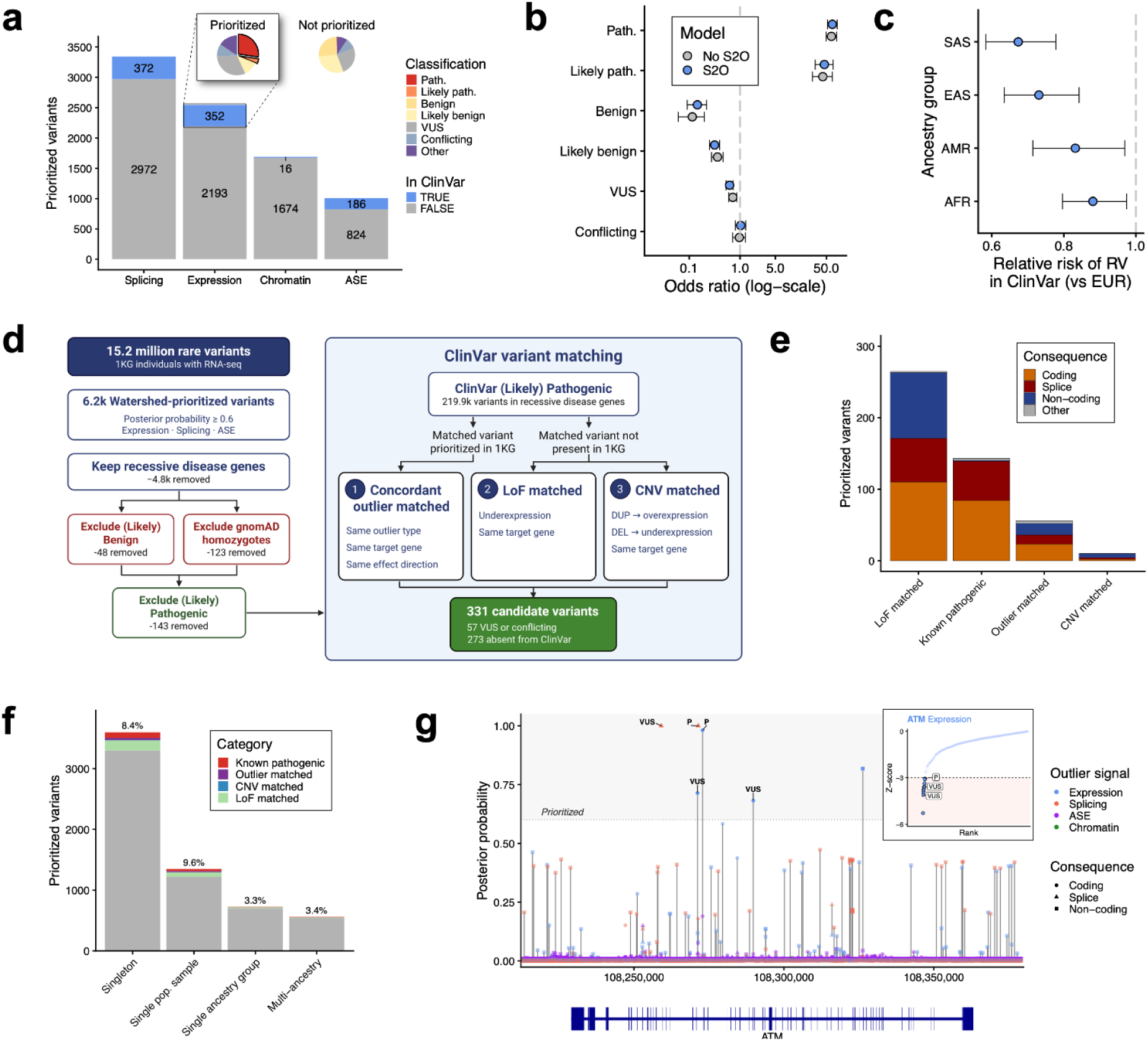
Clinical relevance and population specificity of prioritized rare variants in the 1KG. a) ClinVar representation among Watershed-DL prioritized variants. Inset: ClinVar classifications of prioritized vs. non-prioritized variants in the expression model. b) Enrichment of ClinVar classifications at prioritized vs. non-prioritized variants for Watershed-DL and the baseline Watershed model model without S2O predictions. c) ClinVar enrichment among prioritized variants by ancestry group (chromatin accessibility excluded due to incomplete ancestry representation). d) Nomination of candidate pathogenic variants in recessive disease genes by matching to ClinVar P/LP variants with concordant molecular effects. e) Candidate variants per matching strategy (concordant outlier, LoF, CNV) and known P/LP variants. f) Population specificity of prioritized variants across 30X sr-WGS 1KG samples (singleton, single population, single ancestry group, or multiple ancestry groups), stratified by known P/LP, candidate (panel D), or neither. g) Prioritized variants at the *ATM* locus with ClinVar classifications.

ClinVar is further limited by the European-ancestry bias in clinical variant submissions, resulting in reduced representation of variants from non-European ancestries ^33^. Excluding chromatin accessibility, where not all ancestry groups are represented, 45.20% of prioritized variants were identified only in donors from the five African (AFR) populations in AFGR. The remainder were distributed across the East Asian (EAS), South Asian (SAS), European (EUR), and Admixed American (AMR) ancestry groups (Figure S5B). Prioritized variants from donors of non-European ancestries were less likely to be represented in ClinVar than those from European ancestries (Figure 3C).

To address the under-ascertainment of non-coding variants and variants from donors of non-European ancestries in ClinVar, we developed a molecular effect matching approach to identify disease-relevant candidates among prioritized variants lacking a pathogenic classification. This approach is based on the hypothesis that variants producing a similar molecular effect on a gene as a known pathogenic variant may have similar phenotypic consequences. Among variants prioritized for known recessive disease genes, we first excluded those annotated as benign or likely benign in ClinVar or observed in a homozygous state in gnomAD (v4.1), as homozygous occurrence provides evidence against pathogenicity for variants underlying recessive disease. We then matched the remaining 1,112 prioritized variants, excluding known P/LP variants, to P/LP ClinVar variants using three complementary strategies. The first, *concordant outlier matching*, where prioritized variants have the same molecular outlier as an observed P/LP variant. The second, *LoF matching*, where prioritized variants with underexpression outliers are in the same gene as a known P/LP nonsense or frameshift variant. The third, *CNV matching*, where prioritized variants are in the same gene as a P/LP CNV and with gains matched to overexpression and losses to underexpression (Figure 3D). Together, these matches identified 331 candidate recessive disease alleles, including 57 variants currently classified as VUSs or with conflicting classifications in ClinVar (Figure 3E, Table S5).

Among variants prioritized for gene-level outliers, most were singletons across the 3,202 individuals with 30X sr-WGS in the 1KG (n = 3,599), followed by variants observed in a single population sample (n = 1,349), a single continental ancestry group (n = 724), and multiple ancestry groups (n = 560). Known P/LP variants and candidates identified through molecular effect matching were most concentrated among singletons and single-population variants, underscoring the need for disease allele discovery in ancestrally diverse cohorts (Figure 3F).

The clinical relevance of prioritized variants is further illustrated at *ATM*, an established ataxia-telangiectasia disease gene ^34^, where six variants were prioritized for underexpression and/or splicing outliers, including two P/LP variants and three VUSs (Figure 3G). One VUS was prioritized for an sOutlier and strongly predicted to disrupt the adjacent splice donor (SpliceAI donor loss Δ = 0.71; Pangolin splice loss Δ = 0.76). Another, a missense variant, was prioritized for an underexpression outlier with modest predicted splicing and missense pathogenicity effects (SpliceAI donor gain Δ = 0.16; Pangolin splice gain Δ = 0.17; AlphaMissense = 0.21). The third VUS, a missense variant with high predicted pathogenicity (AlphaMissense = 0.98) but no predicted splicing effect (SpliceAI and Pangolin Δ = 0.00), was prioritized for an underexpression outlier in an individual who also carried an intronic variant predicted to create a cryptic splice donor (SpliceAI donor gain Δ = 0.50; Pangolin splice gain Δ = 0.30). Although Watershed-DL can prioritize more than one variant for a given outlier, the intronic variant received a higher posterior (0.82 vs. 0.68), suggesting it better explains the observed underexpression. Together, this illustrates how Watershed-DL can prioritize variants for clinical interpretation based on both their predicted mechanism and observed molecular effect.

### Watershed-DL prioritizes causal and candidate diagnostic variants in the GREGoR rare disease cohort

We assessed the utility of Watershed-DL for rare disease diagnosis using data from 677 individuals in the GREGoR Consortium (release U15) with matched whole blood RNA-seq and 30X whole genome sequencing. GREGoR enrolls individuals with suspected Mendelian disease, most of whom remain undiagnosed despite previous clinical genetic testing ^26^. This provided an opportunity to benchmark Watershed-DL against known diagnostic variants in solved cases and identify candidate diagnoses in unsolved cases. The cohort included 311 probands, of whom 38 were solved, 27 were partially solved, and 246 remained unsolved (Figure 4A).

**Figure 4:**
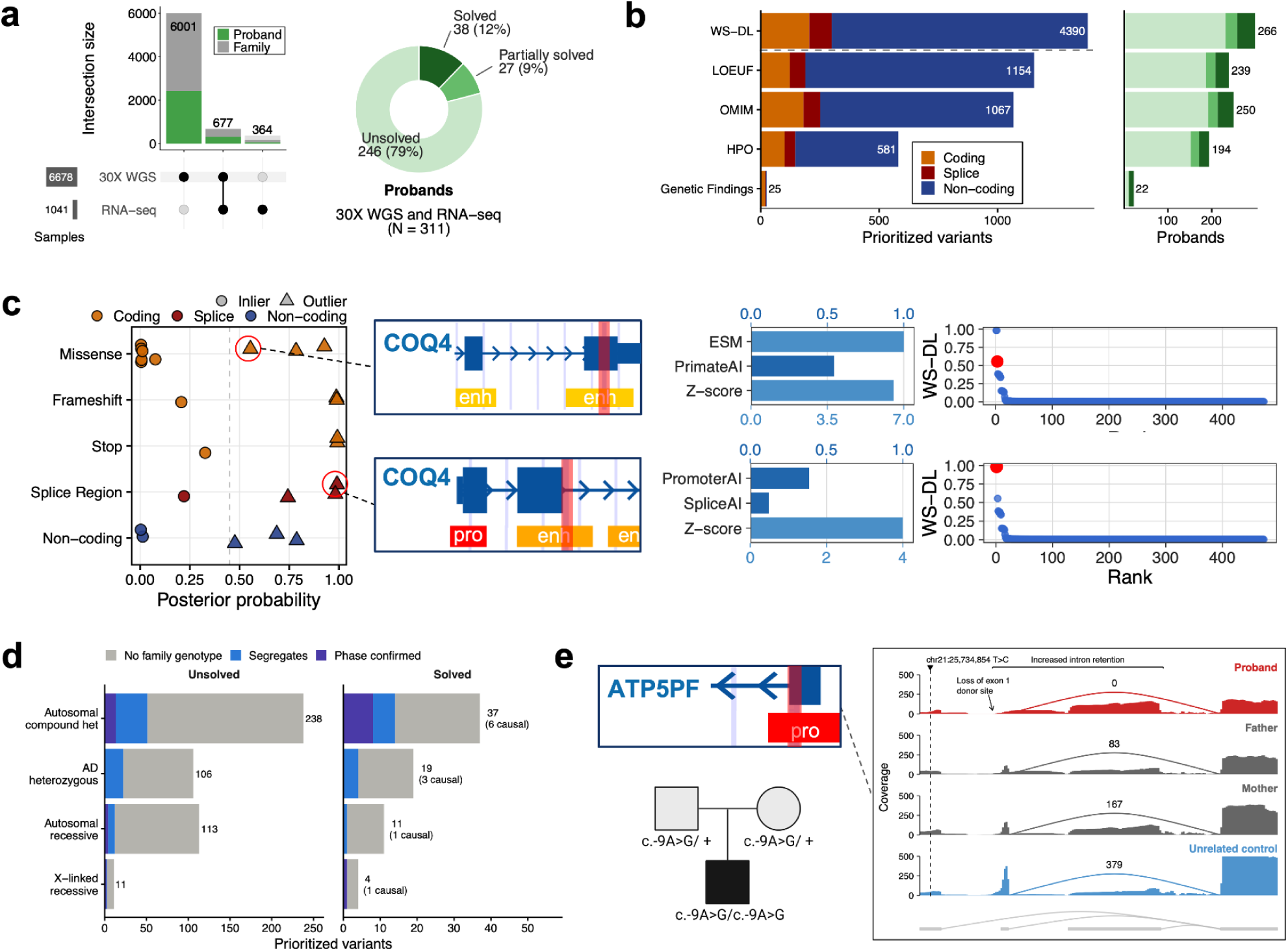
Prioritization of causal and candidate variants in GREGoR probands. a) Overlap of GREGoR individuals with whole blood RNA-seq and 30X sr-WGS. b) Number of Watershed-DL prioritized variants in probands (PP ≥ 0.45), with filters to clinically relevant categories: OMIM disease genes, constrained genes (LOEUF ≤ 0.45), genes matching patient HPO terms, and prior curation. c) Watershed-DL posterior probabilities for 24 diagnostic variants in solved GREGoR probands, stratified by variant consequence, with shape indicating eOutlier status. Cutouts: Two prioritized compound heterozygous variants in a solved proband in COQ4, with S2O features, expression outlier z-scores, and Watershed-DL scores ranked across all scored variants for COQ4. d) Variants prioritized by Watershed-DL in GREGoR probands filtered by inheritance and zygosity criteria, and colored by phenotype segregation status. “Phase confirmed” indicates both parents were genotyped, and alleles were inherited *in trans*. “Segregates” indicates that the variant segregates with affected status in at least one family member. “No family genotype” indicates that no family members were sequenced. e) RNA-seq coverage at *ATP5PF* in an unsolved GREGoR proband carrying a Watershed-DL prioritized homozygous 5′ UTR variant. Coverage is shown for the proband, both heterozygous carrier parents, and an unrelated control.

We trained Watershed-DL to predict eOutliers using a set of S2O predictions which included PromoterAI, SpliceAI, APARENT2, AlphaMissense, ESM-v1, and PrimateAI. Watershed-DL outperformed the genome annotation baseline in the GREGoR RNA-seq cohort (PR-AUC = 0.061 vs. 0.024; Figure S6A) and feature weights were comparable to the 1KG expression model (Figure S6B). For rare disease variant prioritization, we used less stringent outlier thresholds than in the 1KG analysis (posterior probability ≥ 0.45; |z-score| ≥ 2.5). Watershed-DL prioritized 9,324 eOutlier-associated variants across all GREGoR participants, which included 4,390 in probands. Of these, 1,154 were further supported by LOEUF constraint, 1,067 by OMIM disease association, and 581 by HPO phenotype matching, and 25 matched curated variants of interest in the GREGoR *Genetics Findings* table across solved and unsolved cases (Figure 4B).

To assess the sensitivity of Watershed-DL for recovering diagnostic variants, we examined posterior probabilities for diagnostic variants restricted to solved cases in the GREGoR cohort. As the *Genetics Findings* table includes all curated variants, not only those relevant to a patient’s phenotypes, we selected diagnostic variants for each solved proband based on annotated phenotype contribution and mode of inheritance: one variant for autosomal dominant conditions, and either one homozygous variant or two compound heterozygous variants for autosomal recessive conditions (see Methods). This method selected 24 diagnostic variants across 20 probands. Of these, 13 were prioritized in Watershed-DL across 10 probands, which included all diagnostic variants associated with eOutliers in whole blood (Figure 4C). The 11 diagnostic variants not prioritized by Watershed-DL comprised 6 missense variants, 2 intronic variants predicted as donor gain by SpliceAI, and 3 LOFTEE ^1^ loss-of-function variants, none of which were associated with eOutliers in whole blood. In a previously solved proband presenting with spastic diplegia and motor delays, Watershed-DL prioritized two compound heterozygous SNVs in *COQ4* that were previously reported to explain the proband’s phenotype ^35^. These included a maternally inherited splice region variant (SpliceAI donor gain Δ = 0.12, PromoterAI: 0.386) marked as likely pathogenic in the GREGoR *Genetic Findings* table ^36^, and a paternally inherited missense variant (ESM-v1: −7.496, PrimateAI: 0.544, AlphaMissense: 0.296, CADD: 29.8) marked as pathogenic.

To identify Watershed-DL prioritized variants consistent with rare disease inheritance, we filtered prioritized variants by population frequency, phenotype segregation, and phase confirmation where parental sequencing data were available (Table S6; see Methods). In solved probands, inheritance filtering retained 11 out of 13 prioritized diagnostic variants (Figure 4D). The two variants that were not retained by inheritance filtering were each SNVs that formed a compound heterozygous pair *in trans* with an unscored structural variant, and both SNVs were excluded from the autosomal dominant inheritance mode by population frequency criteria. In unsolved probands, inheritance filtering identified 451 candidate variant-gene pairs in 115 probands, including 106 heterozygous candidates for autosomal dominant inheritance, 238 compound heterozygous candidates and 113 homozygous candidates for autosomal recessive inheritance, and 11 candidates for X-linked recessive inheritance (Figure 4D).

Among these, we identified a diagnostic variant in *PPP1R12A* (ENST00000450142.7:c.737A>C) in an unsolved female proband presenting with semilobar holoprosencephaly, seizures, hearing loss, blindness, and scoliosis. The variant is annotated as missense and has an ambiguous prediction from AlphaMissense (score = 0.461). One report that classifies the variant as likely pathogenic exists in ClinVar (RCV005250607.1) but without associated condition or assessment criteria. *PPP1R12A* frameshift and nonsense variants have previously been shown to cause a new syndrome associating brain malformations (in XX and a subset of XY individuals) with urogenital features resulting from impaired testicular development (in XY), termed GUBS (OMIM #618820) ^37^. The proband’s presentation was consistent with the brain malformation features of this syndrome, but because she is an XX female, the absence of genital phenotype was not informative and a diagnosis of GUBS was not suspected prior to molecular testing. The research team had identified the *PPP1R12A* variant as a candidate, but several arguments had prevented reporting into the GREGoR *Genetics Findings* table. All variants reported in Hughes *et al*. were clear stop-gains and *de novo* in affected individuals. Here, the variant is annotated as missense, and examination of LRS reads showed that it did not arise on a maternal haplotype and the father’s sample was not available for comparison (Figure S7A). Watershed-DL prioritized this variant because it was associated with a strong underexpression outlier (z = −3.99), in spite of modest SpliceAI scores for donor loss (Δ = 0.11) and acceptor loss (Δ = 0.06). Consistent with a splicing effect, FRASER 2.0 identified a strong splicing outlier (p < 0.001), and RNA-seq showed skipping of exon 5 in the proband resulting in a frameshift (Figure S7B). The same exon-skipping junction was detected at low levels (∼2% of reads) in the proband’s mother, who does not carry the variant, suggesting that the variant increases usage of a naturally occurring minor isoform rather than creating a novel splice junction. Together, these findings indicate that c.737A>C acts as a missense spliceogenic variant, a class of missense variants that cause disease by disrupting RNA splicing rather than by altering the protein sequence ^38^. By integrating S2O predictions with observed molecular outliers, Watershed-DL revealed a loss-of-function mechanism not captured by the variant’s existing classification, and the variant was confirmed as diagnostic for the proband.

As Watershed-DL is a variant-centric approach that does not rely on prior gene-phenotype relationships, it can also prioritize variants in candidate novel disease genes. In another unsolved proband, we identified a homozygous 5’ UTR variant in *ATP5PF* (ENST00000284971.8:c.-9A>G) that is associated with an underexpression outlier (z = −4.60) and predicted by SpliceAI to cause donor loss (Δ = 0.59). The variant lies within a ∼10.8 Mb region of homozygosity in the proband (chr21 q21.1-21.3) identified by CMA. In this region the parents had an IBS0 count of 12 out of 24,450 sites (0.05%), but were not related genome-wide (Sommalier relatedness 0.04), suggesting a shared haplotype on this region from a common ancestor. Both parents are heterozygous carriers of the prioritized variant, consistent with autosomal recessive inheritance and homozygosity by descent. The variant has no homozygous carriers in gnomAD (v4.1) or All of Us, and *ATP5PF* carries no homozygous loss-of-function variants in gnomAD. RNA-seq shows increased intron retention in the proband and, to a lesser degree, in both carrier parents (Figure 4E). The alternate allele was present in 100% of the proband’s expressed transcripts, but only in 5% and 20% of transcripts in the mother and father, consistent with nonsense mediated decay. The proband presents with spastic quadriplegia, multifocal epilepsy, global developmental delay, dysmorphic features, feeding difficulties, and vision loss. Although *ATP5PF* has no established disease association, biallelic variants in *ATP5ME*, which encodes another subunit of mitochondrial complex V, have been associated with complex V deficiency and a broad range of clinical manifestations, including spasticity, seizures, encephalopathy, feeding difficulties, contractures, hearing loss and optic atrophy ^39^. Similarly, pathogenic variants in other structural subunits of complex V have been linked to autosomal recessive disorders characterized by neurological and neuromuscular dysfunction, encephalopathy, and metabolic abnormalities, with variable disease severity and systemic involvement ^40–45^. Additionally, studies in *ATP14*, the yeast homologue of *ATP5PF*, found that Atp14p depletion severely impairs complex V assembly and ATP synthesis ^46^, while *ATP14* knockout disrupts ATP synthase integrity and function ^47^. Together, these findings make *ATP5PF* a strong candidate disease gene for the proband.

## Discussion

Rare variant prioritization remains a challenging problem with relevant clinical applications in rare disease diagnosis and personalized medicine. Existing prioritization methods typically rely on either molecular outlier signals or sequence-based predictions without integrating these complementary sources of evidence. Here, we present Watershed-DL, an extensible framework that jointly models S2O predictions and personal multi-omic outliers to prioritize rare variants that alter diverse mechanisms such as gene regulation, splicing, and protein function. Watershed-DL improved prioritization of rare variants underlying expression, splicing, and chromatin accessibility outliers relative to both a genome annotation model with S2O predictions, and an outlier-based model without S2O predictions. These gains were greatest for non-coding and non-canonical splice-site variants, for which S2O predictions can provide evidence that is otherwise limited in conventional annotation frameworks. As S2O models continue to improve and multi-omic profiling expands to larger and more diverse cohorts, Watershed-DL offers a scalable framework for interpreting rare variant effects with the flexibility to incorporate improved S2O predictors and new molecular phenotypes as they become available.

Watershed-DL provides interpretable weights which give insight into specific molecular effects of prioritized rare variants and the overall utility of variant annotations to each outlier signal. S2O predictors from matched molecular phenotypes received the largest weights for expression, splicing, and chromatin accessibility, indicating that the model learns biologically meaningful relationships between predicted and observed molecular effects. The contribution of S2O predictions was less pronounced for ASE, where a matched S2O model was not available. In contrast, S2O predictions provided substantial additional evidence for caOutliers, with peak-level modeling extending variant prioritization to distal and intergenic regulatory regions that are often excluded from gene-centric analyses. Together, these results demonstrate that S2O predictions are most informative when they provide variant-level evidence that complements the molecular phenotype observed in an individual, particularly where conventional annotations provide limited evidence of variant function.

Integrating multi-omic outlier signals with S2O annotations enabled the interpretation of rare variants in both self-reported healthy individuals and patients with rare genetic diseases. In the 1KG cohort, Watershed-DL prioritized functional variation in known recessive disease genes, including ancestry-specific alleles absent from current clinical databases. These variants may represent novel recessive disease alleles, disease modifiers, or risk alleles whose clinical significance has yet to be established. In the GREGoR cohort, Watershed-DL prioritized known diagnostic variants in solved cases, including all diagnostic variants associated with eOutliers in whole blood. When combined with evaluation of allele frequency, inheritance patterns and patient phenotypes, it also identified a diagnostic variant in *PPP1R12A* and nominated a candidate gene, *ATP5PF*, not previously associated with disease. Since prioritization does not require genes to have an established disease association, Watershed-DL can also provide functional evidence in novel disease genes harboring rare variants that meet diagnostic criteria. This is particularly relevant for variants outside canonical coding and splice regions, which remain underrepresented in clinical databases and are often overlooked by current diagnostic pipelines.

Several limitations should be considered when interpreting Watershed-DL prioritized variants. Posterior probabilities reflect the combined strength of evidence that a rare variant affects a molecular phenotype, but do not establish that the variant causes the observed effect. A prioritized variant may co-occur with an outlier driven by another variant, an environmental factor, or technical variation, so establishing causality requires experimental validation. In the 1KG cohort, candidate recessive alleles were identified in heterozygous carriers, and the observed molecular effects may not reflect biallelic disruption. In particular, the extent to which partial changes in expression cause disease may be gene-or allele-specific, requiring observation of these variants in affected individuals, either in a homozygous state or *in trans* with a second pathogenic allele. In addition, Watershed-DL does not model relationships between variants and individual traits, and prioritized variants may occur in genes that are not relevant to a patient’s phenotypes. Candidate diagnostic variants identified in GREGoR remain variants of uncertain significance until supported by additional evidence consistent with current RNA-seq diagnostic guidelines ^32^, and may require additional functional or genetic testing specific to the disease or inheritance mode. More broadly, Watershed-DL can only detect effects on genes and regulatory elements that are active in the assayed tissue. Structural variants are also not modeled in the current framework, although approaches for integrating molecular outliers with structural variation have been described ^25^.

Extending Watershed-DL across diverse tissues and cohorts, together with continued improvements in S2O models, will broaden its ability to connect predicted and observed molecular effects to rare variant function. As multi-omic profiling becomes increasingly integrated into clinical genetic testing, Watershed-DL provides a foundation for the systematic variant interpretation needed to enable rare disease diagnosis and delivery of precision medicine.

## Online Methods

### RNA-seq data

RNA-seq data from AFGR ^28^ and MAGE ^29^ were combined to form the 1KG cohort. Lymphoblastoid cell line RNA-seq data for 599 samples from the AFGR cohort were obtained as previously described ^28^. Lymphoblastoid cell line RNA-seq data for 731 samples from the MAGE cohort were downloaded from the Sequence Read Archive at accession PRJNA851328 ^29^. 166 samples from the MKK population of AFGR were excluded due to insufficient rare variant coverage in the VCF, leaving a total of 1,164 RNA-seq samples from 1,030 individuals. Reads were trimmed using Cutadapt ^48^ and aligned to GRCh38 using STAR (v2.7.11b) in two-pass mode with the --waspOutputMode flag. The GENCODE v44 reference annotation was used for alignment. Unless otherwise stated, all analyses were restricted to protein-coding genes. Whole-blood RNA-seq data from 1,041 participants in the GREGoR Consortium (release U12, accession phs003047.v5.p4) were generated at five sites: University of California, Irvine (UCI; n = 534), Stanford University (n = 273), Illumina (n = 93), the Broad Institute (n = 92) and Baylor College of Medicine (n = 49). Participants came from 489 families and included 459 probands. In total, 499 participants were affected (459 probands and 40 family members), 499 were unaffected family members, 24 were possibly affected family members, and 19 were family members with unknown affected status. Library preparation protocols differed between sites, but all libraries were sequenced paired-end. Samples with <75% uniquely mapped reads were excluded.

RNA-seq alignment files, provided in BAM or CRAM format, were converted to FASTQ using SAMtools ^49^. CRAM files were decoded using the GRCh38 full analysis set plus decoy and HLA sequences ^50,51^. Adapters were trimmed with Cutadapt ^48^ and read quality was assessed with FastQC ^52^ before and after trimming. Reads were then shuffled with SeqKit ^53^ for STAR ^54^ compatibility and realigned to a shared human reference (Ensembl v47 ^55^ and GRCh38 primary assembly ^50,51^). Samples with <75% uniquely mapped reads were excluded. Gene-level read counts were obtained for each sample with featureCounts from exon features summarised to genes, counting read pairs from primary alignments only. TPM values were computed from these counts using each gene’s union exon length.

### ATAC-seq data

ATAC-seq data for 100 samples from the AFGR cohort were obtained as previously described ^28^. ATAC-seq data for 100 samples from the Great British population were downloaded from the European Nucleotide Archive (ENA) under accession ERP110508 ^30^. Joint peak calling was done by downsampling the FASTQs to 3 million reads per sample and pooling as previously described ^56^. Three pseudoreplicates of pooled FASTQs were generated and MACS2 peak calling was done using the ENCODE DCC ATAC-seq pipeline available at https://github.com/encode-dcc/atac-seq-pipeline.

### Genotype quality control in the 1000 Genomes cohort

30X short-read whole genome sequencing for samples in the 1KG were obtained from the International Genome Sample Resource (ISGR) ^57^. Genotype quality control was performed using bcftools (v1.21) ^58^. Multi-allelic sites were first split using bcftools norm -m -any. Genotypes with read depth < 10 or genotype quality < 20 were excluded. On the autosomes, variants with missingness > 0.1 or a Hardy-Weinberg equilibrium (HWE) P < 1 × 10⁻⁶ in any super-population were excluded. On the X chromosome, variants with missingness > 0.1 in either XX or XY samples, or with HWE P < 1 × 10⁻⁶ among XX individuals in any super-population were excluded. The pseudoautosomal regions (PAR) were excluded from the X chromosome. Finally, non-variable sites (minor allele count = 0) were removed from all chromosomes. 166 samples from the AFGR Maasai (MKK) subpopulation were excluded from the VCF due to high site missingness.

### Genetic principal components in the 1000 Genomes cohort

Genetic principal components were calculated in the 1KG cohort using PLINK (v2.0) ^59^. Regions with high linkage disequilibrium in hg38 were extracted from plinkQC ^60^ and excluded. For principal component estimation, biallelic autosomal SNVs with minor allele frequency > 5% were retained, and LD pruning was done using –indep-pairwise with a window size of 1Mb, step size of 50, and r^2^ threshold of 0.05.

### Multi-omics outlier calling

#### Gene expression outlier calling in the 1000 Genomes cohort

Gene expression was quantified as transcripts per million (TPM) using RNASeQC (v2.4.3) ^61^. TPM values were log2-transformed (log2(TPM + 2)) and then standardized for each gene to a mean of 0 and a standard deviation of 1. Lowly expressed genes, defined as those with TPM < 0.1 in at least 20% of samples, were excluded. To ensure that the leading principal components (PCs) were not driven by a few samples, we removed samples with highly divergent expression profiles using a sample-network approach implemented in the WGCNA R package ^62^. Pairwise Pearson correlations between all samples were computed across genes and transformed into a signed adjacency matrix (adjacency = 0.5 + 0.5 x correlation). The connectivity of each sample, defined as the sum of its adjacencies to all other samples, was calculated and standardized to a z-score across samples. Samples with a standardized connectivity below −3 were considered global outliers and removed.

Hidden factors were then estimated using the PCAtools (v2.18) R package ^63^ on the filtered expression matrix, with genes centered and scaled. Missing values were imputed using k-nearest neighbors (impute R package ^64^) prior to PCA. A total of 45 expression PCs were retained as selected by the Marchenko-Pastur method. To obtain expression residuals, these expression PCs, the top 5 genotype principal components, batch, RNA Integrity Number (RIN), population, sex, and cohort were removed based on a linear regression model. Expression residuals were then re-scaled to obtain corrected z-scores for each individual per gene.

#### Gene expression outlier calling in the GREGoR cross-consortium cohort

Expression outliers were called on the pooled matrix of 1,041 RNA-seq samples. Lowly expressed protein-coding genes, defined as those with TPM < 1 in more than 20% of samples, were excluded, as were genes with transcription start sites within the major histocompatibility complex (MHC; chr6:28,477,797–33,448,354). TPM values were then log2-transformed (log2(TPM + 2)) and standardized for each gene to a mean of 0 and a standard deviation of 1. Hidden factors were estimated as described for the 1KG data, with the number of components set using the Gavish-Donoho optimal hard threshold for unknown noise. No explicit covariates were included, and the leading principal component explained 50.4% of the variance.

#### Allele-specific expression outlier calling in the 1000 Genomes cohort

Heterozygous sites for each sample were extracted from the VCF using bcftools (v1.21) ^58^. Haplotype-level ASE data were generated using phASER (v1.1.1) ^65^, with a minimum mapping quality of 255 (uniquely mapped reads in STAR) and a minimum base quality of 10. Indels were excluded, and phASER blacklists were applied to remove sites and haplotypes prone to mapping artifacts. Haplotypes were phased genome-wide using the most common haplotype phase (--gw_phase_method 0), based on the phase information in the VCF.

To obtain gene-level ASE, GENCODE v44 annotations were collapsed so that each gene was represented by a single gene model of merged exons across its transcripts. Haplotypic counts were then aggregated per gene using phASER Gene AE, which sums read counts supporting each haplotype across the phased heterozygous variants within the gene’s exons and reports the log2 allelic fold change (aFC) between the two haplotypes. Genes with missing values in at least 80% of samples were removed, and the remaining log2 aFCs were scaled for each gene to a mean of 0 and a standard deviation of 1. As described for gene expression outlier calling, samples with highly divergent ASE profiles were excluded prior to PCA, missing values were imputed, and hidden factors were estimated, retaining 65 ASE PCs selected by the Marchenko-Pastur method. ASE residuals were obtained by residualizing these PCs and the same covariates used for expression, then re-scaling for each gene to obtain corrected z-scores.

#### Splicing outlier calling in the 1000 Genomes cohort

Splicing outliers were called using FRASER 2.0 ^22^, which models aberrant splicing using an intron Jaccard index. FRASER 2.0 was run using the FRASER R/Bioconductor package (v2.4.5) within the DROP pipeline (v1.4.0) ^66^, with GENCODE v44 to annotate introns. Introns were retained with ≥ 20 split reads in one sample and ≥ 10 reads in 75%, matching DROP defaults. Low variance introns were removed using a minimum deltaPsi threshold of 0.3, which requires at least one sample to differ from the intron’s mean Jaccard index by greater than 0.3. The default HGNC IDs were replaced with Ensembl IDs during the annotation step. The default DROP pipeline minimum read count and expression settings were used, filtering to a ≥ 20 split reads in one sample and ≥ 10 reads in 75% of samples. Sample covariation was removed with a PCA-based denoising autoencoder, and the encoding dimension was selected using the optimal hard threshold procedure. For each sample, a gene-level p-value was defined as the minimum Holm-adjusted p-value across all introns in that gene. Finally, splicing outliers (p < 0.0027) with a delta-Jaccard index below 0.3 were excluded to ensure significant outliers reflect large splicing changes.

#### Splicing outlier calling in the GREGoR cohort

Splicing outliers were called using FRASER 2.0 ^22^ (v2.6.0) in the GREGoR UC Irvine RNA-seq cohort using GENCODE v47 basic annotation and the harmonised STAR alignments. 537 UCI whole-blood samples were fitted as a UCI-only cohort. Introns were kept with ≥ 20 split reads in one sample and ≥ 10 reads in 75% of samples, matching DROP pipeline defaults. The encoding dimension was selected with the optimal hard threshold procedure using the PCA implementation.

#### Chromatin accessibility outlier calling in the 1000 Genome cohort

Chromatin accessibility outliers were called using EpiOut (v0.0.1) ^67^ on the joint peak set from all 200 AFGR and GBR ATAC-seq samples described above. Read counts per peak and sample were generated with EpiCount, with the default merging of nearby peaks disabled. EpiOut was then run on the resulting count table to model accessibility and obtain a p-value, observed and expected read count, and log2 fold change for each peak in each sample. To remove outliers with small effect sizes, the EpiOut effect size and coverage criteria were applied, excluding observations with p < 0.01 that had an absolute log2 fold change below 0.5, or that had both observed and expected read counts below 50. Peak-level p-values were then converted to z-scores, signed by the direction of the log2 fold change. A p-value threshold of p < 0.0027 was used to define chromatin accessibility outliers.

### Rare variant filtering

For each cohort, variants were left-aligned and normalized, multiallelic sites were split into biallelic records, and spanning deletions were removed using bcftools (v1.21) ^58^. Sites with FILTER = PASS or FILTER = “.” were retained. Sites with ≥10% of genotypes missing were removed in the 1KG. In GREGoR, a higher threshold of ≥90% missing genotypes was used to avoid removing rare variants, as the cross-consortium U15 release VCF was merged from separately joint-called VCFs across 89 and 588 samples respectively. Variants were then restricted to those with a cohort-level allele frequency ≤5%, and insertions and deletions longer than 30 bp were removed. Finally, variants with allele frequency >1% in any of the AFR, AMR, ASJ, EAS, SAS or NFE populations of gnomAD (v4.1) ^68^ were removed. Variants absent from gnomAD were retained.

### Rare variant annotation

#### VEP annotations

To annotate rare variant consequences, VEP annotations were generated by running Ensembl VEP (v110.1, GRCh38, Gencode v44) ^69^, using a window size of 10kb to define upstream and downstream consequences. Loss-of-function annotations were generated using the LOFTEE (GRCh38) plugin for VEP ^1^. The UTRAnnotator VEP plugin ^70^ was used to annotate 5’ UTR consequences such as premature start codon gain or loss. Annotated consequences were retained for transcripts with transcript support level 1 or 2, Ensembl canonical transcripts, and regulatory features.

For Watershed-DL, VEP consequences were each represented as categorical variables. For region-level aggregation, a per-column maximum over all rare variants in the region was used. For analyses requiring a single consequence per rare variant-gene pair, each variant was assigned its most severe VEP consequence according to the following hierarchy: coding [frameshift > stop > start > missense > other coding], splice [splice donor > splice acceptor > splice region > other splice], non-coding [5′ UTR > 3′ UTR > regulatory > intron > upstream > downstream > intergenic > other non-coding]. For the chromatin accessibility model, variants were assigned their most severe non-coding consequence only.

#### Polyadenylation site annotation

Polyadenylation sites were annotated using PolyA-DB v3.2 ^71,72^ after lifting over from hg19 to hg38 using the liftOver function from the rtracklayer R package ^73^. Variant proximity to polyadenylation sites was represented as a binary annotation indicating whether the variant is within 50bp of an annotated polyadenylation site. The absolute distance to the nearest polyadenylation site was also included as an annotation. PolyA-DB genes were matched to the corresponding VEP gene annotation and outlier gene.

#### Enhancer-to-gene and CTCF loop annotations

To link distal regulatory regions to genes, enhancer-gene links and promoter-anchored CTCF loops were compiled from ENCODE-rE2G predictions and ChIA-PET data, respectively, in the lymphoblastoid cell line GM12878. ENCODE-rE2G predictions (accession ENCFF128MZW) and CTCF ChIA-PET loops (accession ENCFF780PGS) were obtained from the ENCODE portal. Each CTCF loop was split into its two anchors, and anchors overlapping an EPDnew v6 promoter (hg38; https://epd.expasy.org/epd/) were assigned to the corresponding gene. Variants were annotated with E2G or CTCF links for a given gene if they fell within an enhancer or opposite loop anchor linked to that gene.

#### ClinVar annotations

ClinVar classifications were obtained from the variant summary file (variant_summary.txt.gz) downloaded from the NCBI FTP site (https://ftp.ncbi.nlm.nih.gov/pub/clinvar/tab_delimited/) on September 6, 2026, and restricted to GRCh38 coordinates. Classifications were harmonized by merging subcategories into their parent terms (e.g., low-penetrance and VUS subcategories). Variants with multiple classifications for the same gene and phenotype were collapsed to a single classification according to the following priority: pathogenic > benign > likely pathogenic > likely benign > uncertain significance > conflicting classifications > not provided. Molecular consequences for each variant were obtained from the MC field of the ClinVar GRCh38 VCF (release 2026-09-05; https://ftp.ncbi.nlm.nih.gov/pub/clinvar/vcf_GRCh38/). For gene-level outliers, ClinVar classifications were assigned to prioritized variants only if the ClinVar gene matched the outlier gene.

### Sequence-to-omics predictions

#### Predictions of chromatin accessibility

Chromatin accessibility predictions were annotated for all SNVs and short indels (≤ 30bp) using ChromBPNet ^7^. Pre-trained, bias-corrected ChromBPNet models trained on ATAC-seq samples from the AFGR cohort ^28^ were used, including 30 models (6 population-specific training sets, 5 folds each). Variants were restricted to those falling exactly within joint-called ATAC peaks. For each variant and model, the predicted effect on accessibility was calculated as a log-fold allele change between reference and alternate alleles. The average absolute log fold change across all 30 models was used as the final annotation.

#### Predictions of gene expression

RNA-seq read coverage in LCLs was predicted for rare SNVs and short indels (≤ 30bp) in the 1KG using Flashzoi ^27^, an optimized implementation of Borzoi ^5^, referred to here as Flashzoi-RNA. A single replicate of the pre-trained Flashzoi model, trained only on human data, was downloaded from Huggingface (https://huggingface.co/collections/johahi/flashzoi-models). Predictions were made using the full 524kb input sequence, with predicted RNA-seq read coverage output at 32 base-pair resolution. Scores were averaged across the GM12878 lymphoblastoid cell line RNA-seq tracks from ENCODE (ENCFF463TDW, ENCFF279MVV).

For the Watershed-DL transcriptomic model, all SNVs within 60kb of GENCODE v44 protein-coding genes were scored. The input window was centered at the gene’s transcription start site, and coverage was summed across the gene’s exons. Variant effects were quantified as the log-fold change in summed predicted coverage between reference and alternate alleles (logSED). For the chromatin accessibility model, all SNVs within ATAC-seq peaks were scored, with the 524kb input sequence centered on the variant. Variant effects were quantified as the L2 norm of the log difference in predicted coverage between reference and alternate alleles across the binned output sequence.

#### Prediction of promoter usage

Promoter site usage predictions were annotated with PromoterAI ^6^, which predicts CAGE-seq based on sequence. Pre-computed scores for all SNVs within promoters (+-100bp of TSS) of protein-coding genes annotated in Gencode v39 were downloaded from GitHub (https://github.com/Illumina/PromoterAI). To capture rare variant effects on expression, the PromoterAI model fine-tuned on outlier-associated rare variants in GTEx was used. Scores represent the difference in predicted CAGE-seq reads between reference and alternate alleles averaged across the output window, which includes 4096 nucleotides centered at the variant. Scores range from −1 to 1, with negative values indicating predicted decreases in promoter activity and positive values indicating predicted increases.

#### Predictions of splice site usage

Predicted splicing effects were annotated using SpliceAI ^8^ and Pangolin ^9^. Pre-computed SpliceAI scores for SNVs within canonical GENCODE v24 transcripts, generated using a ± 50bp window around each variant, were downloaded from Illumina BaseSpace (https://basespace.illumina.com/s/otSPW8hnhaZR). Short indels (<30bp) in the 1KG cohort were scored separately with a pretrained SpliceAI model using the VEP SpliceAI plugin, using the distributed GENCODE v24 configuration and a ± 50bp scoring window around each variant. For each variant, delta scores for acceptor gain, acceptor loss, donor gain, and donor loss were annotated, defined as the change in predicted acceptor or donor probability between reference and alternate alleles. Each variant was assigned the SpliceAI score for the gene matching its outlier signal by HGNC ID.

Pre-trained Pangolin models were obtained from GitHub (https://github.com/tkzeng/Pangolin), which includes three models per tissue for heart, liver, brain, and testis. All SNVs and short indels (≤ 30bp) within GENCODE v44 gene bodies were scored using a 500bp scoring window around each variant. Splice site gain and loss scores were annotated as the largest gain and loss within the window across all tissues.

#### Predictions of transcript polyadenylation

Predicted effects on polyadenylation were annotated using APARENT2 ^10^. APARENT2 predicts the relative usage of a polyadenylation site, quantified by the proportion of RNA-seq transcripts cleaved at that site. Pre-computed scores for all SNVs near polyadenylation sites annotated in PolyADB-v3 ^71^ were obtained from CADD (v1.7) ^74^. Variant scores represent the log-odds ratio of predicted polyadenylation site usage between reference and alternate allele. Odds ratios were included as the final annotation.

#### Predictions of missense variant pathogenicity

Missense variants were annotated for predicted pathogenicity using AlphaMissense ^13^, ESM-1v ^11^, and PrimateAI ^12^. AlphaMissense scores were annotated using the VEP plugin ^69^, which uses pre-computed scores for all possible missense SNVs based on canonical protein-coding transcripts in GENCODE v32. AlphaMissense was included as a binary annotation representing whether a variant is predicted as likely pathogenic, defined by the standard threshold of predicted pathogenicity ≥ 0.564. PrimateAI (v0.2) scores were annotated using the VEP plugin and included as raw predicted scores, which range from 0 to 1 with higher scores indicating greater predicted pathogenicity. ESM-1v scores for all possible SNVs were obtained from CADD (v1.7) and included as raw scores. ESM-1v scores represent the log-likelihood ratio of predicted deleteriousness for reference and alternate amino acids, with more negative scores indicating greater predicted deleteriousness.

### Watershed-DL model training and validation in the 1KG

#### Assigning variants to genes and peaks

To match variant annotations to gene-level and peak-level outlier signals, variants were first assigned to genes using GENCODE (v44) annotations or to peaks using the joint peak calls. GENCODE (v44) genes were filtered to protein-coding genes.

For gene-level outlier signals (expression, splicing, ASE), variants were assigned to a gene if they fell within a 10kb window around the gene body as defined by GENCODE (v44). VEP consequences were filtered to match the annotated GENCODE gene. For S2O predictions linked to a specific gene, the prediction for the assigned gene was used where available, and otherwise predictions were averaged across genes. For model training and evaluation, each annotation was summarized across all variants assigned to a given individual-gene pair, using the minimum or maximum so that the most severe annotation was retained (Table S1,S2). After training, Watershed-DL was applied to predict variant-level posterior probabilities, using each individual’s observed outlier status for the assigned gene.

For chromatin accessibility outliers, variants were assigned to a peak if they overlapped the peak region. Similar to the gene-level model, each annotation was summarized across all variants within a peak for each individual during training and evaluation, using the minimum or maximum value (Table S2). After training, Watershed-DL was applied to predict variant-level posterior probabilities, using each individual’s observed outlier status for the assigned peak.

#### Multi-omic outlier preprocessing

Prior to training Watershed-DL, AFGR samples from the Maasai population (MKK) were excluded as described above (17 samples from ATAC-seq and 166 from RNA-seq). RNA-seq and ATAC-seq samples not in the sr-WGS were also excluded, removing 1 sample from the RNA-seq and 9 samples from the ATAC-seq. Since 134 samples from AFGR and MAGE corresponded to the same donors, z-scores from replicate samples were averaged for each transcriptomic outlier signal. Across gene-level and peak-level outliers, a standard p-value threshold of p < 0.0027 (|z| ≥ 3) was used to define outliers. Expression and ASE p-values were signed by direction of effect. Splicing and chromatin accessibility p-values were unsigned. The Watershed-DL model thresholds unsigned p-values into inlier (p > 0.0027) and outlier (p < 0.0027) categories, and thresholds signed p-values into underlier (p < 0, |p| < 0.0027), overlier (p > 0, |p| < 0.0027), and inlier (|p| > 0.0027) categories, with independent model weights defining impact on functional status for each category. For each outlier signal, global outlier individuals were removed, defined as individuals carrying a higher outlier burden as defined by the mean plus a standard deviation threshold (σ ≥1.5 for expression and chromatin, σ ≥2 for ASE and splicing, Table S4). After filtering, outlier p-values were joined to the gene-level or peak-level aggregated features by individual and region.

#### Training the Watershed-DL model

The Watershed-DL model was trained with exact inference using the distributed R package (https://github.com/nicolerg/WatershedR). Models were trained on region-level summarized annotations, which reduces noise during training caused by individuals carrying multiple rare variants with mixed functional status in a given region. N2-pairs, defined as two individuals with the same set of variants in a given region (gene or peak), were excluded from training and held out for evaluation. For regions where three or more individuals carry identical rare variants, annotations and outlier signals were averaged across individuals prior to training. The default Dirichlet prior (C = 10) and a standardized L2 penalty (λ = 0.001) were used for all models and outlier signals. Models were trained until convergence, defined as a change of less than 1 × 10^−8^ between iterations. To assess the impact of S2O annotations and multi-omic outliers in Watershed-DL, two baseline models were also trained: a Watershed model without S2O predictions, and a genome annotation model without outlier signals.

#### Validation on variant-matched pairs

Following previous Watershed work ^15,23–25^, performance was evaluated on held-out N2 pairs. For each pair, posterior probabilities were computed for one individual using their variant annotations and outlier status, and this posterior was used to predict the outlier status of the second individual. This approach provides an unbiased proxy of model performance when true functional status is unobserved. Performance was summarized as the area under the precision-recall curve across posterior thresholds.

#### Imputation at the variant level

After training, posterior probabilities were computed across all individuals and variants within each cohort, resulting in 1 prediction per individual, gene, and variant. In the 1KG, variants were considered prioritized if they had PP ≥ 0.6, following prior work ^25^, and were carried by at least one individual with an outlier in the assigned gene or peak (|p| < 0.0027).

### Identification of candidate recessive disease alleles in the 1KG

To identify candidate recessive disease alleles, prioritized variants were restricted to recessive disease genes curated from ClinGen, OMIM, and ACMG. ClinGen genes were defined as those with a haploinsufficiency score of 30 (gene associated with autosomal recessive phenotype) in the ClinGen Dosage Sensitivity curation list (accessed September 9, 2026) ^75^. OMIM genes were defined as those with at least one phenotype annotated with autosomal recessive inheritance in the OMIM genemap2 file (accessed September 2, 2023) ^76^. ACMG genes were defined as those with autosomal recessive inheritance in the ACMG Secondary Findings list (v3.3) ^77^.

Within these genes, prioritized variants were compared with pathogenic or likely pathogenic (P/LP) variants in ClinVar. Variants already classified as P/LP or as benign or likely benign in ClinVar were excluded, as were variants observed in a homozygous state in gnomAD (v4.1). Variants absent from gnomAD were retained. For each remaining variant, the direction of effect was defined as the sign of its mean outlier z-score across carriers. Each variant was then matched to ClinVar P/LP variants using three criteria: (1) concordant outlier match, in which a P/LP variant in the same gene was also prioritized in 1KG and its carriers showed outliers in the same direction; (2) LoF match, in which the variant was prioritized for an underexpression outlier and the gene contained at least one P/LP nonsense or frameshift variant; and (3) CNV match, in which a P/LP CNV overlapped the gene, with copy number gains matched to overexpression outliers and losses matched to underexpression outliers. Variants could satisfy more than one criterion. For analyses requiring a single match type per variant, matches were prioritized as follows: concordant outlier > CNV > LoF. All matches are reported in Table S5.

### Watershed-DL analysis in GREGoR rare disease cohort

#### Assigning variants to genes

To match variant annotations to gene-level and peak-level outlier signals, variants were first assigned to genes using GENCODE (v47) annotations, and genes were filtered to protein-coding genes. For expression outliers, variants were assigned to a gene if they fell within a 10kb window around the gene body as described in the 1KG. VEP consequences were filtered to the annotated GENCODE gene, and S2O predictions linked to a specific gene were filtered to the assigned gene were available, and averaged across genes where a match was unavailable. For training and evaluation, annotations were summarized across all variants assigned to a given individual-gene pair using the minimum or maximum so that the most severe annotation was retained (Table S3). After training, variant-level posterior probabilities were computed as described for the 1KG.

#### Watershed-DL model training and variant scoring

For Watershed-DL training and variant scoring in the GREGoR cohort, the GREGoR U15 joint-called VCFs were downloaded from AnVIL (release U15, accession phs003047.v5.p4) ^26^. Among samples in the VCF, 677 overlapped with the U15 RNA-seq data. Following the protocol described for 1KG, global outlier individuals (σ >1.5) and individuals carrying no outliers were removed from the training data (Table S4). Expression outliers were defined as |p| < 0.0027, and Watershed-DL was trained using the default Dirichlet prior of C = 10 and an L2 penalty of λ = 0.001. N2 pairs were held out during training and AUPRC was evaluated on this held-out set. After training, variant-level posterior probabilities were computed over variant-gene pairs in all individuals, excluding global outliers and individuals with no outliers. For variant-level scoring, a higher outlier threshold of |p| < 0.01 was used to increase sensitivity to sub-threshold outliers in disease-relevant genes.

#### Candidate variants in conserved and disease-relevant genes

To assess disease relevance for prioritized variants, known disease genes were annotated using OMIM ^76^, filtering to genes where at least one phenotype has a known molecular basis. Genes were matched to prioritized variant-gene pairs by Ensembl gene ID.

Patient phenotype relevance was assessed by annotating prioritized variants with each proband’s HPO terms from the GREGoR U14 phenotype table, downloaded from AnVIL. For each prioritized variant-gene pair, the proband’s HPO terms were matched to HPO terms annotated to the gene using the pyhpo library (v4.0.0; https://pypi.org/project/pyhpo/), including both direct matches and descendant terms. Gene symbols in the HPO database were mapped to Ensembl gene IDs using HGNC.

Prioritized variant-gene pairs were also annotated with loss-of-function tolerance using the gnomAD (v2.1.1) loss-of-function observed/expected upper bound fraction (LOEUF) ^1^, with genes with LOEUF ≤ 0.45 considered loss-of-function intolerant.

#### Prioritization of previously curated variants

To analyze Watershed-DL prioritization of previously curated variants, the GREGoR *Genetics Findings* table (version U15) was downloaded from AnVIL. Variant-gene pairs scored in probands were matched to curated variants based on the variant position, alternate allele, and Ensembl gene ID.

To assess recovery of diagnostic variants in solved probands, curated findings were restricted to probands marked as Solved. For probands with findings in more than one gene, only genes with the strongest phenotype contribution were retained, ranked as Full > Partial > Uncertain. Multiple genes were retained only for probands with more than one gene marked as Partial and no gene marked Full. For findings annotated as autosomal recessive, probands were required to carry a homozygous or hemizygous variant, or at least two heterozygous variants in the annotated gene.

#### Inheritance filtering criteria

Prioritized variants were evaluated for family segregation and phase under several inheritance models: autosomal dominant, autosomal recessive (homozygous and compound heterozygous), and X-linked recessive (homozygous, hemizygous, and compound heterozygous) (Table S6). Genotypes for probands and sequenced relatives were extracted from the GREGoR joint-called VCF (version U15) using bcftools (v1.21) ^58^, and the sex and affected status of relatives were obtained from the GREGoR *Participant* table (version U14). Population-level allele frequencies for all ancestry groups, homozygote counts, and hemizygote counts were extracted from gnomAD (v4.1). Population-level allele frequency and homozygote count was downloaded from the All of Us (AoU) public data browser. Variants absent from either resource were assigned an allele count of 0. Under the autosomal dominant model, variants were required to have an allele frequency ≤ 1 × 10^−6^ in both gnomAD and AoU. Under recessive models, variants were required to have no homozygotes in gnomAD or AoU, and, for X-linked inheritance, allele frequency ≤ 1 × 10^−6^ among males in gnomAD. Variant segregation was assessed based on the affected status, genotype, and sex of each sequenced relative. Relatives with unknown affected status were excluded from filtering criteria. For compound heterozygous variants, phase was considered confirmed when each parent carried exactly one of the two variants. Variants with no testable relatives were retained if they passed population-level allele frequency and count criteria, but were labeled as lacking family genotypes.

## Data and code availability

Code for Watershed-DL is being made available at https://github.com/e-271/Watershed-DL. Watershed-DL posteriors for the 1000 Genomes Project and GREGoR samples are being made available on Zenodo (DOI: 10.5281/zenodo.23426012). RNA-seq and ATAC-seq data from AFGR are available on ENCODE at https://www.encodeproject.org/search/?searchTerm=AFGR&type=Experiment. RNA-seq data from MAGE are available from the SRA under accession PRJNA851328. ATAC-seq data from the GBR population are available from the ENA under accession ERP110508. 30X sr-WGS genotypes for 1000 Genomes samples are available from the IGSR at https://www.internationalgenome.org/data-portal/data-collection/30x-grch38. RNA-seq and sr-WGS data from GREGoR are available in AnVIL (release U15, accession phs003047.v5.p4).

## Acknowledgments

We thank the participants and families of the GREGoR Consortium and the 1000 Genomes Project for their contributions to this research. J.L. and T.M.A. were supported by the National Science Foundation Graduate Research Fellowship Program under Grant No. DGE-2146755. V.R. was supported by the National Human Genome Research Institute of the National Institutes of Health under training grant T32HG000044. The GREGoR Consortium is funded by the National Human Genome Research Institute of the National Institutes of Health, through the following grants: U01HG011744, U01HG011745, U01HG011755, U01HG011758, U01HG011762, and U24HG011746. The content is solely the responsibility of the authors and does not necessarily represent the official views of the National Institutes of Health. Computing for this project was performed on the Stanford SCG Bioinformatics Cluster (RRID:SCR_026876), owned by the Stanford Genomics Bioinformatics Service Center (RRID:SCR_023340) and operated by Stanford Research Computing (RRID:SCR_023413).

## Supplementary Materials

**Figure S1:**
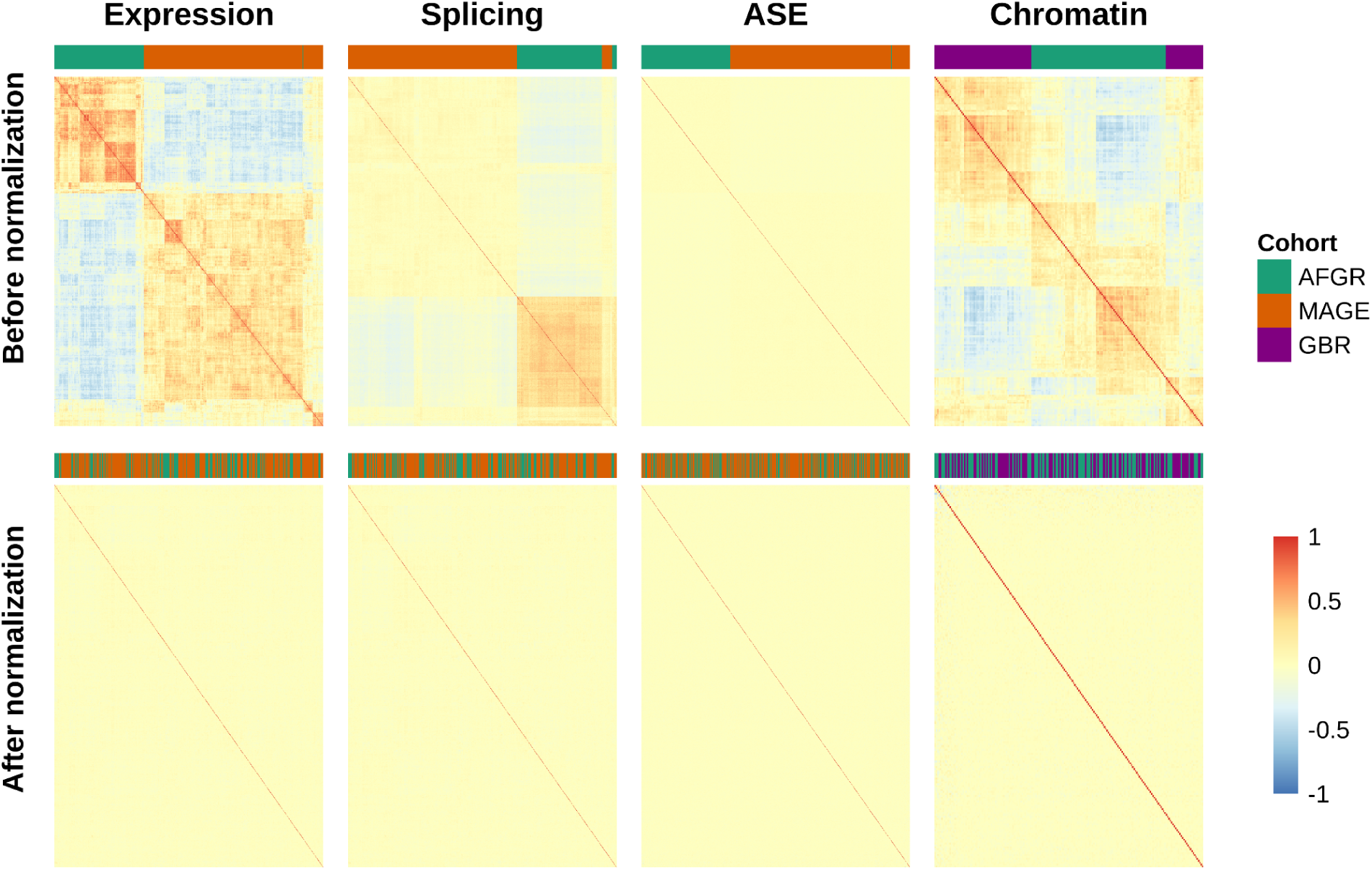
Sample–sample correlations before and after correction for confounders. Heatmaps show pairwise Spearman correlations between samples, computed on the following values for each outlier signal. Expression: centered and scaled log TPM for genes with TPM ≥0.1 in at least 20% of samples, before and after correction for known and hidden factors. Splicing: Intron Jaccard Index for the 15,000 most variable junctions, before and after FRASER 2.0 autoencoder correction. ASE: centered and scaled allelic log2 fold change, before and after correction for known and hidden factors. Chromatin accessibility: centered and scaled log CPM for the 15,000 most variable peaks before correction, and log2 fold change after EpiOut autoencoder correction. Samples corresponding to the same donor from AFGR and MAGE were excluded from visualization.

**Figure S2:**
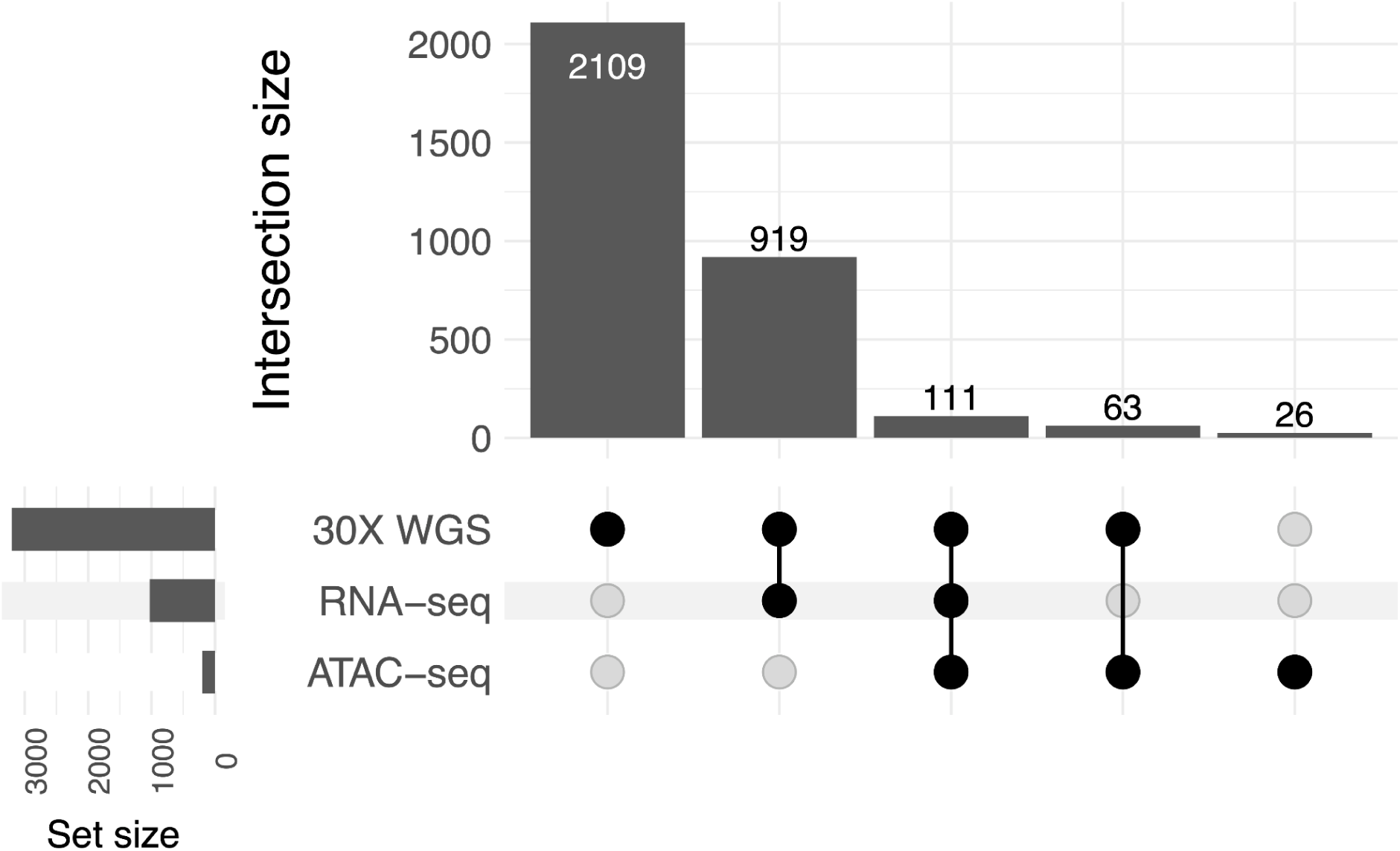
Overlap of samples with RNA-seq, ATAC-seq, and 30X sr-WGS in the 1KG. Upset plot showing the overlap of samples with 30X sr-WGS, RNA-seq, and ATAC-seq in the 1KG. 1,030 samples with sr-WGS and RNA-seq were included in the Watershed-DL model. 174 samples with sr-WGS and ATAC-seq were included in the ATAC only model.

**Figure S3:**
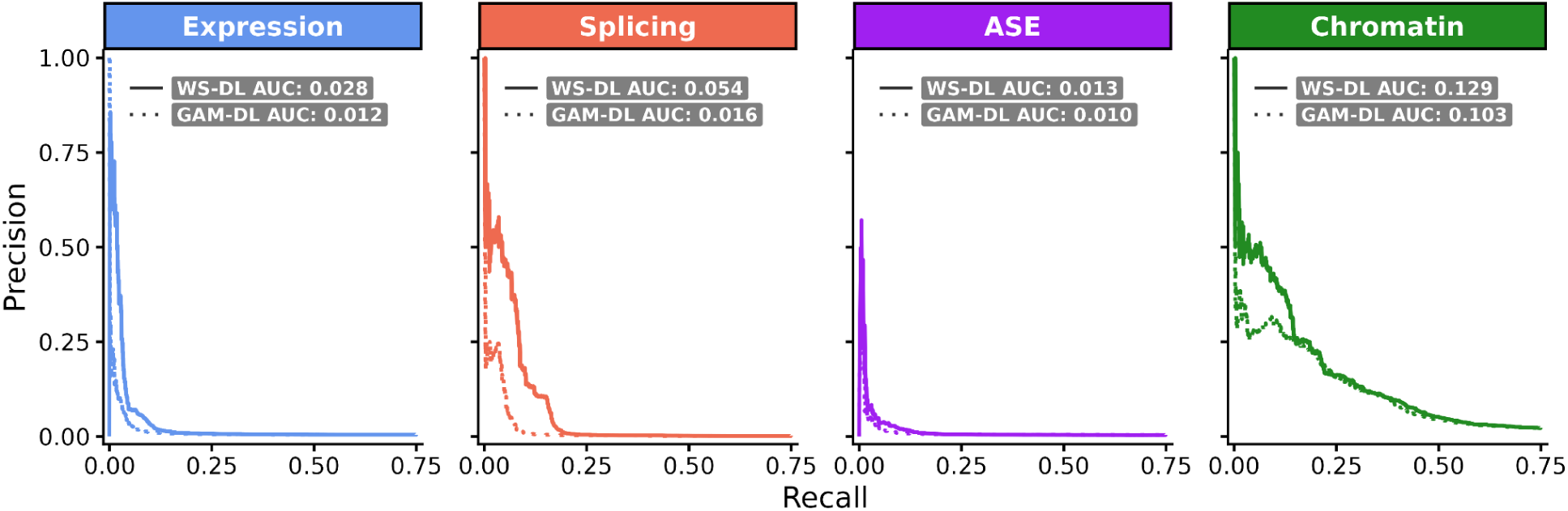
Comparison of precision-recall performance between Watershed-DL and genome annotation model with S2O predictions (GAM-DL). Precision-recall curves on held-out N2 pairs comparing Watershed-DL (solid lines) to a genome annotation model with S2O predictions (dashed lines) across outlier signals in the 1KG.

**Figure S4:**
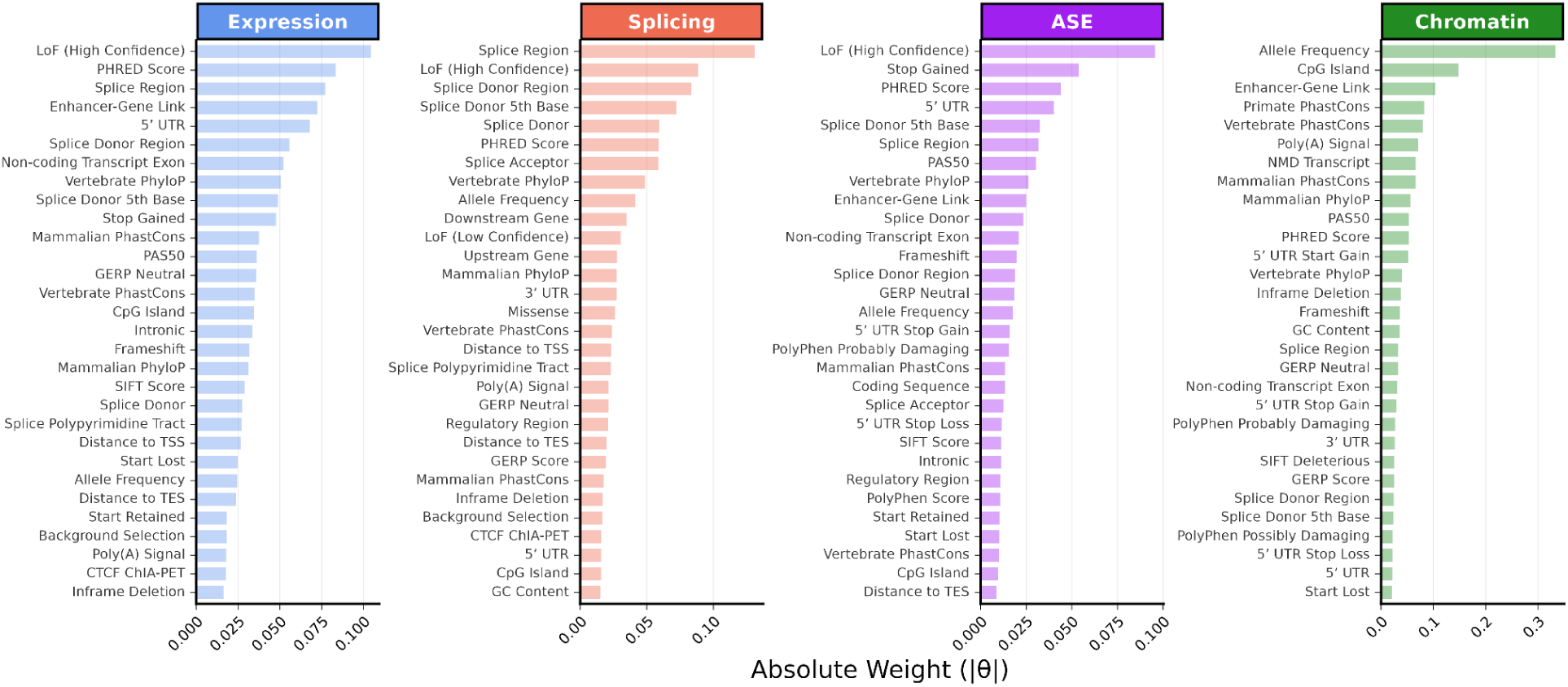
Edge weights connecting genomic annotation features (G) to latent regulatory variables (Z) in the Watershed model without S2O predictions. For each outlier signal, features are ranked by |θ| value representing relative feature informativeness. The top 30 most informative annotations are shown.

**Figure S5:**
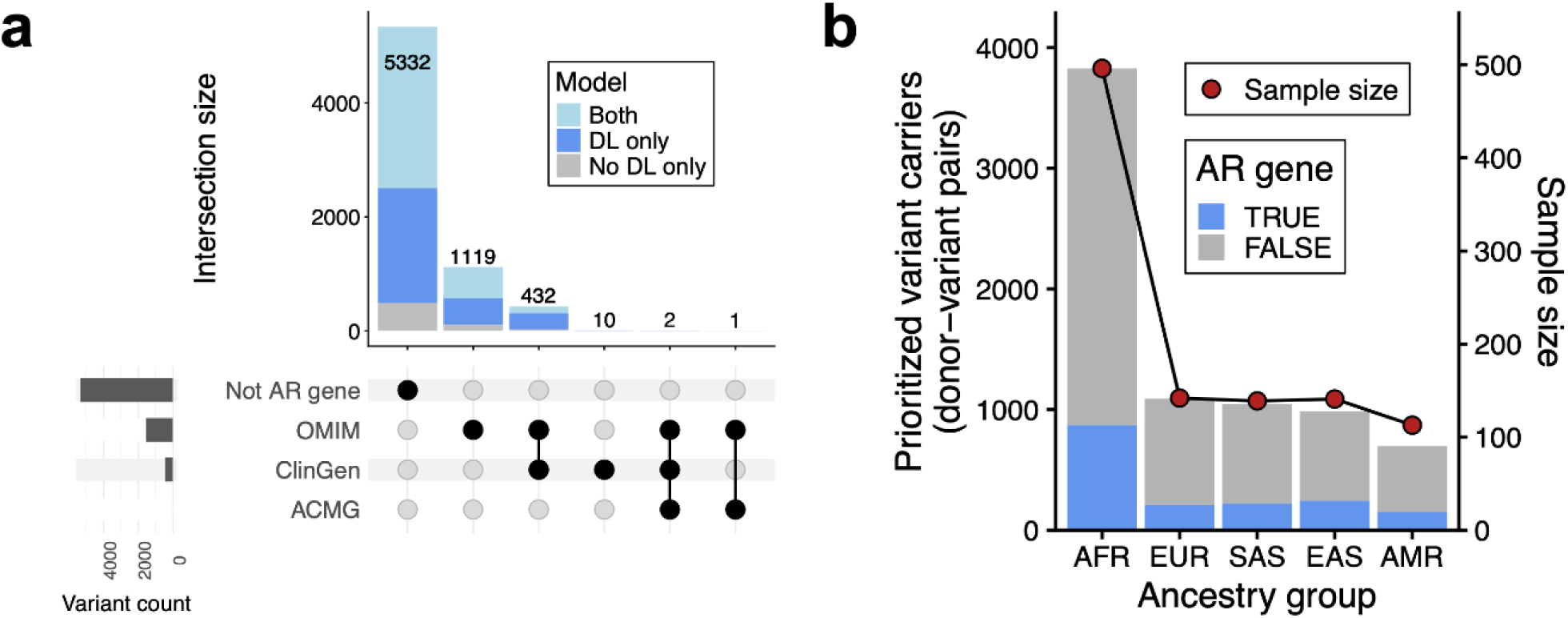
Disease gene overlap and ancestry distribution of Watershed-DL prioritized variants. a) Number of Watershed-DL prioritized variants in genes belonging to recessive disease gene sets from OMIM, ClinGen, and ACMG. b) Ancestry group distribution of prioritized variants from Watershed-DL. Points indicate expected counts based on sample size relative to AFR.

**Figure S6:**
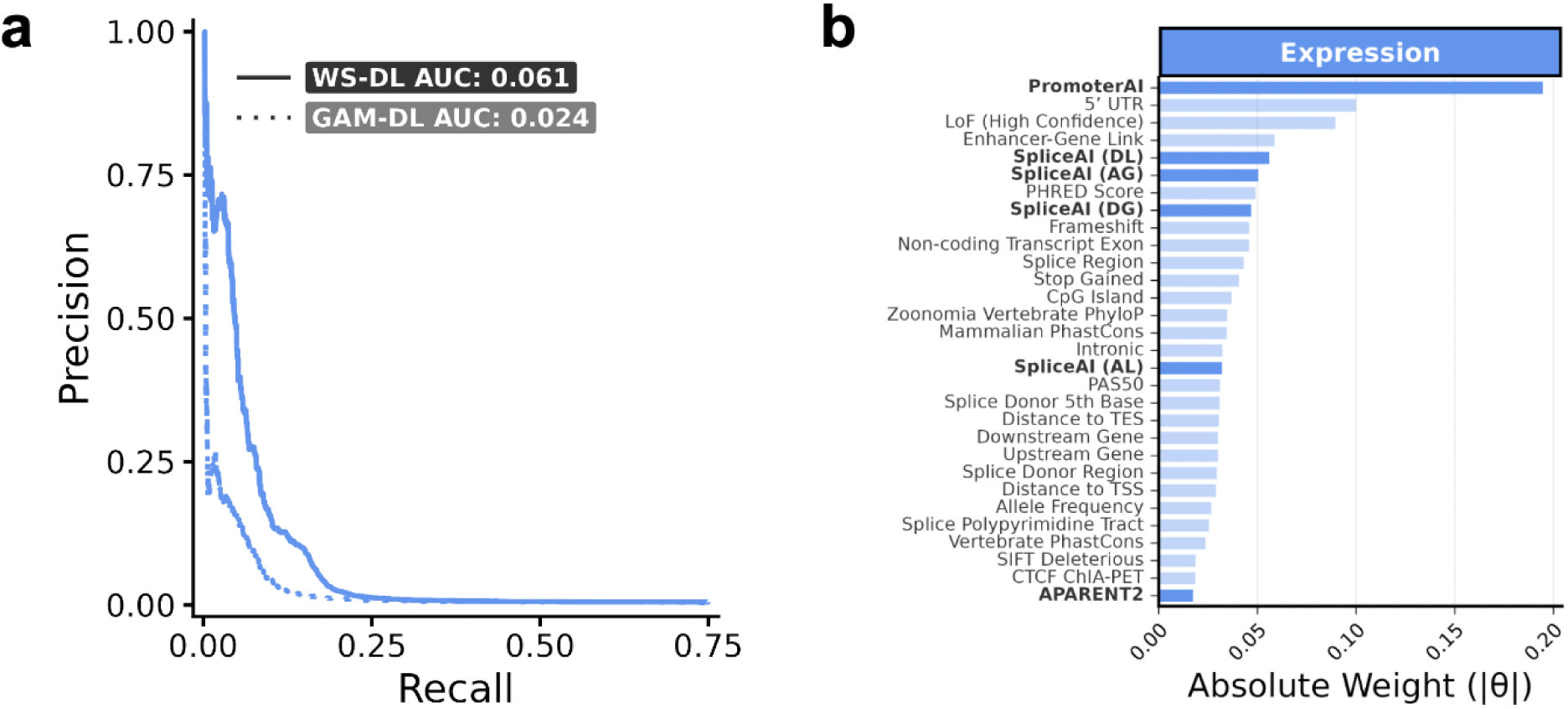
Watershed-DL benchmarks in the GREGoR cohort. a) Precision-recall curves for Watershed-DL (solid line) and a genome annotation model with S2O predictions (GAM-DL) (dashed line) in the GREGoR cohort. b) Edge weights connecting genomic annotation features (G) to the expression latent variable (Z) in the Watershed-DL model trained on GREGoR, ranked by |θ| value. The top 30 most informative annotations are shown.

**Figure S7:**
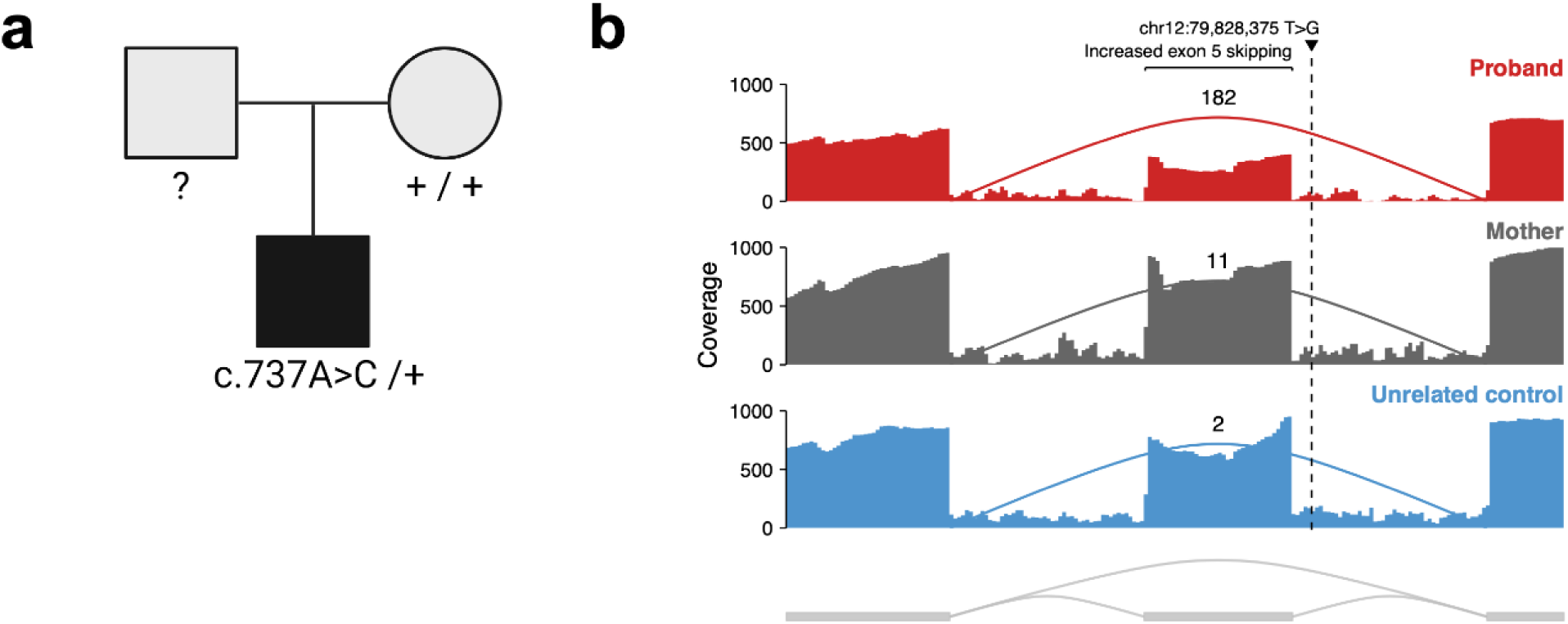
Splicing effect at *PPP1R12A* in a previously undiagnosed proband with semilobar holoprosencephaly. a) Pedigree showing *PPP1R12A* variant segregation. The variant is present in the affected proband and absent in the mother; the father is unavailable for testing. b) Sashimi plots of RNA-seq read coverage across *PPP1R12A* in the proband, the proband’s mother, and an unrelated control. The proband has a junction that skips exon 5, which was also detected at low levels (∼2% of reads) in the proband’s mother.

### Supplementary Tables

**Table S4:** Watershed-DL outlier p-value thresholds and global outlier criteria.

| Cohort | Outlier signal | P-value threshold | Max outlier standard deviation | Max outliers per individual | Global outlier individuals removed | Zero-outlier individuals removed |
| --- | --- | --- | --- | --- | --- | --- |
| 1KG | Expression | 0.0027 | 1.5 | 222 | 59 | 0 |
| 1KG | Splicing | 0.0027 | 2 | 83 | 60 | 0 |
| 1KG | ASE | 0.0027 | 2 | 77 | 19 | 7 |
| 1KG | ATAC | 0.0027 | 1.5 | 1067 | 16 | 2 |
| GREGoR | Expression | 0.0027 | 1.5 | 131 | 79 | 3 |

